# Multiplexed CRISPR-based microfluidic platform for clinical testing of respiratory viruses and SARS-CoV-2 variants

**DOI:** 10.1101/2021.12.14.21267689

**Authors:** N.L. Welch, M. Zhu, C. Hua, J. Weller, M. Ezzaty Mirhashemi, S. Mantena, T.G. Nguyen, B.M. Shaw, C.M. Ackerman, S.G. Thakku, M.W. Tse, J. Kehe, M.R. Bauer, M-M. Uwera, J.S. Eversley, D.A. Bielwaski, G. McGrath, J. Braidt, J. Johnson, F. Cerrato, B.A. Petros, G.L. Gionet, S.K. Jalbert, M.L. Cleary, K.J. Siddle, C.T. Happi, D.T. Hung, M. Springer, B.L. MacInnis, J.E. Lemieux, E. Rosenberg, J.A. Branda, P.C. Blainey, P.C. Sabeti, C Myhrvold

**Author notes:** These authors contributed equally (M. Zhu, C. Hua). These authors supervised this work (J.A. Branda, P.C. Blainey, P.C. Sabeti, C Myhrvold).

## Abstract

The COVID-19 pandemic has demonstrated a clear need for high-throughput, multiplexed, and sensitive assays for detecting SARS-CoV-2 and other respiratory viruses as well as their emerging variants. Here, we present microfluidic CARMEN (mCARMEN), a cost-effective virus and variant detection platform that combines CRISPR-based diagnostics and microfluidics with a streamlined workflow for clinical use. We developed the mCARMEN respiratory virus panel (RVP) and demonstrated its diagnostic-grade performance on 533 patient specimens in an academic setting and then 166 specimens in a clinical setting. We further developed a panel to distinguish 6 SARS-CoV-2 variant lineages, including Delta and Omicron, and evaluated it on 106 patient specimens, with near-perfect concordance to sequencing-based variant classification. Lastly, we implemented a combined Cas13 and Cas12 approach that enables quantitative measurement of viral copies in samples. mCARMEN enables high-throughput surveillance of multiple viruses and variants simultaneously.

## Introduction

COVID-19 has exposed critical gaps in our global infectious disease diagnostic and surveillance capacity^1^. The pandemic rapidly necessitated high-throughput diagnostics to test large populations^2^, yet early diagnostic efforts met technical challenges that cost the United States precious time in its early response^3,4^. Other challenges developed as the pandemic progressed that point towards an additional need for highly multiplexed surveillance technologies. These challenges include the co-circulating human respiratory viruses that cause symptoms similar to COVID-19^5,6^ and emerging SARS-CoV-2 variants of concern (VOCs) with mutations that impact viral fitness and clinical disease prognosis^7,8^.

An ideal diagnostic would also have surveillance capabilities to process hundreds of patient samples simultaneously, detect multiple viruses, differentiate between viral variants, and quantify viral load^9,10^; yet no such test currently exists. As it stands, there is a trade-off between clinically approved high-throughput diagnostics and multiplexed methods in the number of patient samples and/or pathogens tested simultaneously^11–13^. As examples, reverse transcription-quantitative PCR, RT-qPCR is high-throughput by testing at least 88 samples, but for 1-3 analytes at a time^3,14,15^; multiplexed techniques such as, Cepheid Xpert Xpress detects 4 respiratory viruses in up to 16 samples per run^16^, and BioFire detects 22 respiratory pathogens in 1 sample simultaneously^17^. Additionally, these and other clinical diagnostic methods, except ChromaCode^18,19^, do not comprehensively detect SARS-CoV-2 variant mutations^20,21^. Instead, detecting SARS-CoV-2 variants has largely been achieved through next-generation sequencing (NGS)^22,23^, which is time-consuming, expensive, and requires bioinformatic expertise to interpret^7,24–27^.

CRISPR-based diagnostics offer an alternative approach to detecting multiple viruses and variants^28–30^. CRISPR effector proteins Cas12 or Cas13 activate upon CRISPR RNA (crRNA)-target binding which unleashes their collateral cleavage activity on a fluorescent reporter for readout of viral positivity status^31–34^. The crRNA-target binding events are highly specific and altered by the presence of sequence variation. Maximally active crRNA design has been accelerated by machine learning and other computational methods^35^. Nonetheless, most CRISPR diagnostics detect one or two targets per sample^31,36^, which is not sufficient for differential diagnosis via comprehensive microbe or variant identification.

To scale up the capabilities of CRISPR-based diagnostics, we developed Combinatorial Arrayed Reactions for Multiplexed Evaluation of Nucleic acids (CARMEN)^37^ which parallelizes nucleic acid detection (Fig. 1a). The first generation of CARMEN, referred to here as CARMEN v1, could detect 169 human-associated viruses in 8 samples simultaneously. In CARMEN v1, samples and Cas13-crRNA complexes remain separately confined for barcoding and emulsification prior to pairwise droplet combination for detection by fluorescence microscopy. This allows each sample to be tested against every crRNA. CARMEN v1 is a powerful proof-of-concept for multiplexed CRISPR-based detection, but it is difficult to use in a clinical setting given its use of custom-made imaging chips and readout hardware, manually intensive 8-10 hour workflow, and low-throughput sample evaluation.

**Figure 1.**
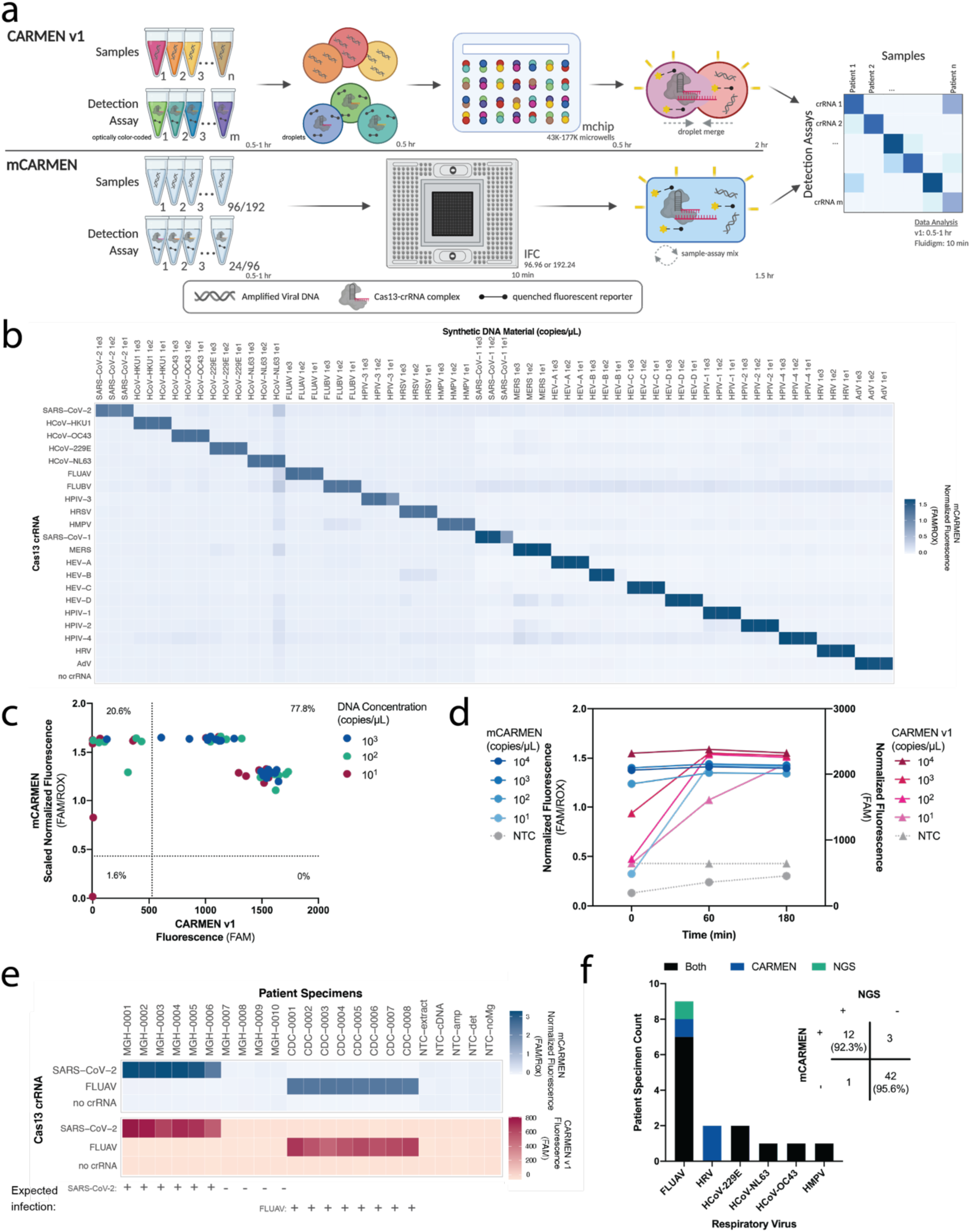
CARMEN implementation on Fluidigm achieves greater sensitivity quicker. **a**, Schematic of CARMEN v1 (top) and mCARMEN (bottom) workflows. **b**, Heatmap showing mCARMEN fluorescent data across 21 human respiratory viruses that were amplified using two separate primer pools. Synthetic DNA fragments were serially diluted from 10^3^-10^1^ copies/μL and added to Q5 amplification master mix. All samples were background subtracted from NTC-noMg negative control. **c**, Concordance between CARMEN v1 and mCARMEN from **b**. Blue: targets at 10^3^ copies/μL; green: targets at 10^2^ copies/μL; red: targets at 10^1^ copies/μL. **d**, Fluorescence kinetics of amplified SARS-CoV-2 DNA gene fragments from 10^4^-10^1^ copies/μL at 0, 60, and 180 minutes post-reaction initiation. Blue: mCARMEN; red: CARMEN v1. **e**, Testing 21 human respiratory virus panel on clinical specimens from 6 SARS-CoV-2 positive, 4 SARS-CoV-2 negative NP swabs, and 8 FLUAV positive specimens, collected prior to Dec. 2019, and 5 no target controls (NTCs). Heatmap shows fluorescent signals from SARS-CoV-2 crRNA, FLUAV crRNA and no crRNA control. Blue: mCARMEN; red: CARMEN v1. **f**, Concordance of mCARMEN and NGS on 58 suspected respiratory virus infected patient specimens collected prior to Dec. 2019. Black: detected by both mCARMEN and NGS; blue: detected by mCARMEN only; green: detected by NGS only. mCARMEN values are shown as normalized fluorescence, FAM signal divided by passive reference dye, ROX, signal at 1 hour post-reaction initiation. CARMEN v1 values are shown as raw fluorescence, FAM signal at 3 hours. NTC, no target control; NTC-extract, no target control taken through extraction, cDNA synthesis, amplification, and detection; NTC-cDNA, no target control taken through cDNA synthesis, amplification, and detection; NTC-amp, no target control taken through amplification and detection; NTC-det, no target control taken through detection; NTC-noMg, no target control expected to have no fluorescent signal due to lack on Mg2+ needed to activate Cas13.

To fulfill the public health need for a clinically relevant surveillance technology that detects multiple viruses and variants quickly, we developed microfluidic CARMEN (mCARMEN). mCARMEN builds on CARMEN v1 and uses commercially available Fluidigm microfluidics and instrumentation^38^. mCARMEN on Fluidigm allows for a faster and less labor-intensive alternative to CARMEN v1 and requires 3x less sample input compared to single analyte RT-qPCR testing. The mCARMEN workflow can process 188 patient samples for up to 23 respiratory viruses and variants in under 5 hours, and costs <$10 USD per sample (Supp. Table 1). In an academic setting, we detected 21 human respiratory viruses by mCARMEN (Supp. Table 2). Then, in a CLIA-certified laboratory at Massachusetts General Hospital (MGH), we validated the condensed respiratory virus panel (RVP), comprised of nine common viruses and a human internal control, RNase P. Further, we developed a SARS-CoV-2 variant identification panel (VIP) which we immediately applied to the detection of Omicron as cases emerged in the Boston area without any changes to the assay. Additionally, we combined Cas13 detection with Cas12 to quantify viral genome copies in samples, yielding results comparable to RT-qPCR. To our knowledge, mCARMEN is the only diagnostic that combines surveillance capabilities into a single technology platform with the ability to test hundreds of samples in a day for multiple respiratory viruses and variants, while also being able to quantify viral genomic copies.

## Results

### CARMEN implementation on Fluidigm for detecting 21 human respiratory viruses

CARMEN v1^37^ is limited by custom instrumentation requirements and labor-intensive protocols which is why we sought to develop a scalable technology that could be broadly implemented. Microfluidic CARMEN (mCARMEN) meets these requirements and eliminates the color-coding and dropletization needs of CARMEN v1 by using commercially available integrated fluidic circuits (IFCs) on the Fluidigm Biomark^38^ (Fluidigm, San Francisco, CA) (Fig. 1a). By leveraging Fluidigm microfluidics, we overcame the need for a custom microscope and chips as well as data analysis expertise, which were required for CARMEN v1. The Fluidigm IFCs use a specific number of assay combinations: 192 samples by 24 detection assays or 96 samples by 96 detection assays which are all spatially separated. After manual IFC loading, the Fluidigm controller moves the samples and detection assays through individual channels on the IFC until they reach the chip reaction chamber, where they are thoroughly mixed. We measure fluorescence on the Fluidigm Biomark with our custom automated protocols that take images of the IFC chip every 5 minutes for 1-3 hours at 37°C (Extended Data Fig. 1a).

In our first implementation of the mCARMEN platform, we designed a panel to detect 21 clinically relevant human respiratory viruses (Supp. Table 2). This includes all viruses covered by BioFire RP2.1 - SARS-CoV-2, four other human-associated coronaviruses and both influenza strains - as well as a few additional illness inducing viruses^39^. To generate maximally active virus-specific crRNAs and PCR primers to detect the 21 viruses, we applied the assay design method ADAPT (<u>A</u>ctivity-informed <u>D</u>esign with <u>A</u>ll-inclusive <u>P</u>atrolling of <u>T</u>argets; described in methods)^35^. We were able to encompass the full genomic diversity of these viral families by including multiple primers, if needed.

We compared the performance of mCARMEN to CARMEN v1 for detecting synthetic DNA fragments recapitulating the 21 viral targets, and found mCARMEN had the same (13 viruses) or better (8 viruses) analytical sensitivity compared with CARMEN v1 (Fig. 1b and 1c, Extended Data Fig. 1b). Both mCARMEN and CARMEN v1 had 100% analytical specificity, but mCARMEN was 100% sensitive to 10^2^ copies/μL and 98.4% sensitive to 10^1^ copies/μl while CARMEN v1 was only 86% and 77.8% sensitive, respectively. Moreover, the mCARMEN reaction rate is accelerated compared with CARMEN v1, resulting in faster initial detection and signal saturation of targets (Fig. 1d, Extended Data Fig. 1c). This is likely due to the higher temperature at reaction initiation for mCARMEN (37°C) than for CARMEN v1 (25°C), and the extensive sample-detection assay mixing that occurs in the mCARMEN IFC, rather than merged droplets mixing by diffusion in CARMEN v1.

We then benchmarked the performance of both CARMEN diagnostics against RT-qPCR^40^ and/or unbiased metagenomic NGS on patient specimens. We obtained a set of 6 SARS-CoV-2-positive, 4 SARS-CoV-2-negative, and 8 influenza A virus (FLUAV)-positive patient specimens for initial testing. mCARMEN and CARMEN v1 had 100% concordance with RT-qPCR, NGS, and each other (Fig. 1e). We also compared performance using two different fluorescent reporters, RNase Alert (IDT, Coralville, IA) and a custom 6-Uracil-FAM (polyU) reporter^34^. We found enhanced sensitivity when using a polyU fluorescent reporter due to LwaCas13a’s preference to cleave at uracils^28,29^ (Extended Data Fig. 2a&b).

Aside from SARS-CoV-2 and influenza viruses, the remaining 19 viruses detectable by mCARMEN lack a recognized gold-standard clinical diagnostic. Thus, we compared mCARMEN to unbiased metagenomic NGS results for the characterization of 58 pre-pandemic unknown samples collected from patients with a presumed upper respiratory infection (Fig. 1f, Supp. Table 3, Extended Data Fig. 2c&d). Both mCARMEN and NGS detected the same respiratory viruses in 12 specimens (7 FLUAV, 2 HCoV-229E, 1 HCoV-NL63, 1 HCoV-OC43, and 1 HMPV), neither detected respiratory viruses in 42 specimens, and they had differing results for 4 specimens, with 93% overall concordance based on an average of ~3 million reads per specimen. Nine of the 12 specimens positive by both methods assembled complete genomes while the remaining 3 assembled partial or no genomes but had >10 reads (2 FLUAV, 1 HMPV). mCARMEN missed 1 virus-positive specimen detected by NGS, a partial FLUAV genome. We found no sequencing reads spanning the mCARMEN amplicon, suggesting degradation was responsible for the result. mCARMEN detected virus in 3 specimens (1 FLUAV, 2 HRV) where NGS did not detect any viral reads. While we cannot rule out false positive results, metagenomic sequencing has been shown to have poor sensitivity for low viral copy samples^6,24,41^.

### Streamlining mCARMEN respiratory virus panel for future clinical use

With a drive towards clinical applications, we aimed to optimize the mCARMEN workflow. To do so, we decreased the manual labor and processing time from >8 hours to <5 hours by implementing automated RNA extraction, using a single-step RNA-to-DNA amplification with 1 primer pool, and reducing the duration of detection readout (Fig. 2a, Extended Data Fig. 3a). We then preliminarily evaluated the optimized workflow on 21 SARS-CoV-2 positive and 8 negative patient specimens, and found greater sensitivity over the original two-step amplification method (Extended Data Fig. 3b).

**Figure 2.**
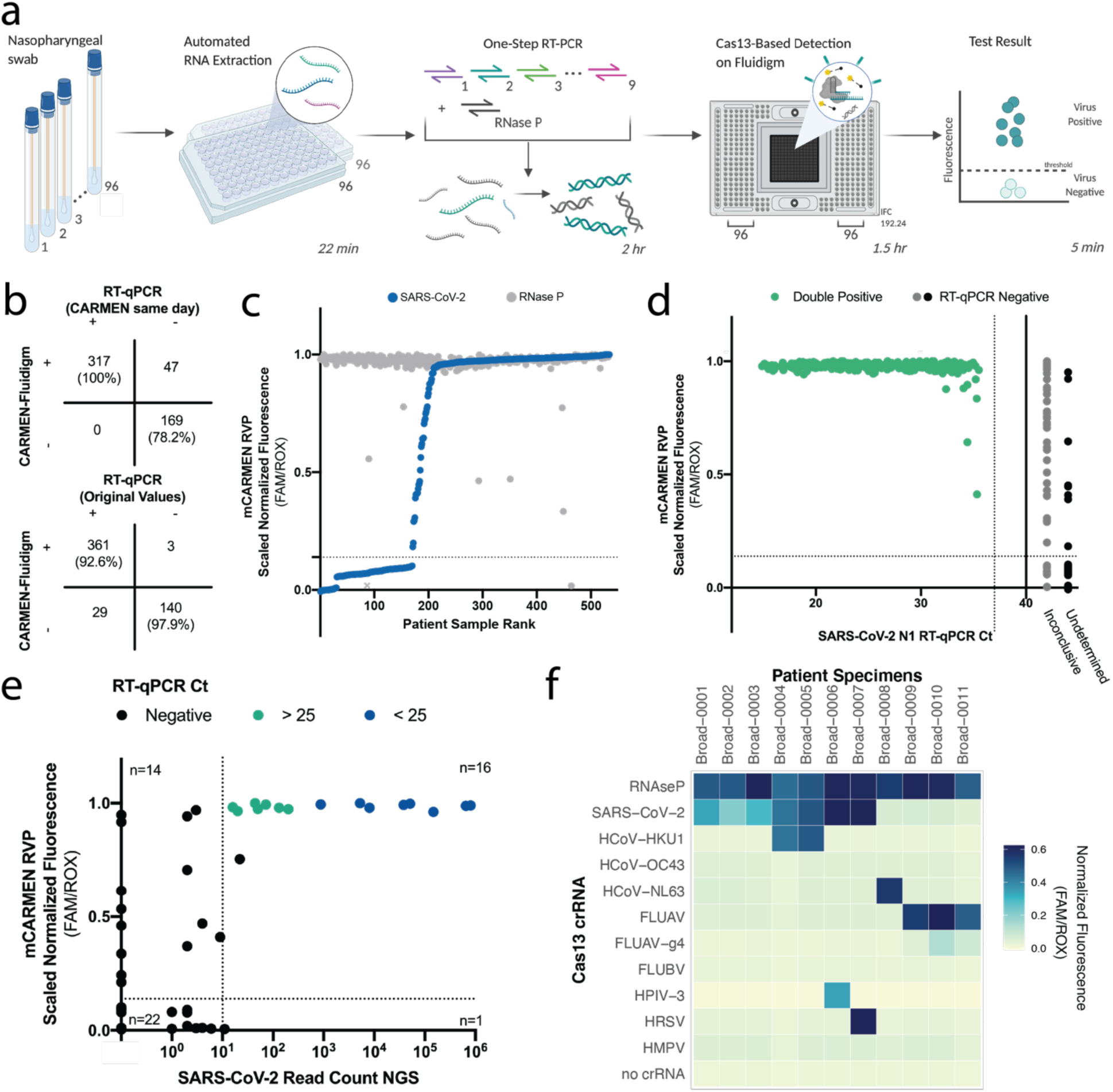
Automated and condensed mCARMEN workflow evaluated on >500 NP swabs. **a**, Schematic of streamlined mCARMEN workflow to test 188 patient specimens for a panel of 9 human respiratory viruses, RVP (SARS-CoV-2, HCoV-HKU1, HCoV-OC43, HCoV-NL63, FLUAV, FLUBV, HPIV-3, HRSV, HMPV) and a human internal control (RNase P). **b**, Concordance of RVP and RT-qPCR results; RT-qPCR results from concurrent testing (top), and RT-qPCR results from original testing (bottom). **c**, Scaled normalized fluorescence at 1 hour for 533 NP swabs ranked by increasing SARS-CoV-2 signal (blue) with the respective RNase P signal (gray). Fluorescence is normalized by dividing the FAM signal by ROX then data are scaled from 0 to 1. NTC-noMg signal as 0 and the maximum normalized fluorescence value at 1 hour as 1. Dashed horizontal line: threshold for RVP positivity. Threshold is calculated by multiplying the NTC-extract fluorescence value by 1.8; NTC-extract: no template control taken through the entire workflow. Gray X represents the single failed sample excluded from concordance calculations and other analysis. **d**, Scatter plot of the scaled normalized fluorescence values from **b** compared to viral Ct values obtained from concurrent testing with the ThermoFisher TaqPath COVID-19 Kit. Green: positive SARS-CoV-2 signal detected by both RVP and RT-qPCR; gray: inconclusive RT-qPCR result indicating one or two of the three technical replicates were undetermined; black: undetermined RT-qPCR result indicating all three technical replicates were negative for SARS-CoV-2. Dashed horizontal lines: threshold for RVP positivity. Dashed vertical line: Ct value of 37 (FDA positivity cutoff). **e**, Concordance of RVP scaled normalized fluorescence values and SARS-CoV-2 read counts by unbiased NGS. Dashed horizontal line: threshold for RVP positivity; dashed vertical line: 10 reads mapped to the SARS-CoV-2 genome by NGS, threshold for NGS positivity. Blue: SARS-CoV-2 N1 RT-qPCR Ct < 25; green: Ct > 25; black: negative by RT-qPCR. **f**, Normalized fluorescence values at 1 hour for 11 patient specimens.

For an end-to-end mCARMEN workflow, we developed software to be used alongside clinical testing to provide patient diagnoses (Extended Data Fig. 6). The software uses the final image at 1 hour post-reaction initiation as input, then automatically validates controls to make 1 of 3 calls: “detected,” “not detected,” or “invalid” for each combination of sample and crRNA.

Lastly, we wanted to condense mCARMEN for focused clinical use and did so by developing a respiratory virus panel (RVP) to detect 9 of the most clinically-relevant viruses (SARS-CoV-2, HCoV-HKU1, HCoV-OC43, HCoV-NL63, FLUAV, FLUBV, HPIV-3, HRSV, and HMPV) and a human internal control, (RNase P). These nine viruses were included on RVP based on if they heavily circulate in the population and have capacity to cause respiratory virus symptoms, while others were excluded if genomic diversity was difficult to account for concisely, such as HRV^42^. We first conducted range-determining limit of detection (LOD) studies for the 9 viruses on mCARMEN RVP in a research laboratory. The preliminary LOD was within the range of 100-1,000 copies/mL for SARS-CoV-2, FLUAV, FLUBV, HCoV-HKU1, HCoV-NL63, HCoV-OC42, and 1,000-20,000 copies/mL for HPIV-3, HMPV, HRSV (Extended Data Figure 5a, Supp. Table 4), with robust performance from the SARS-CoV-2 crRNA as well as all RVP crRNAs in combination (median AUCs of 1 and 0.989, respectively) (Extended Data Figure 5b-g).

To benchmark mCARMEN RVP performance to comparator assay results, we analyzed 390 SARS-CoV-2-positive and 143 negative patient specimens, and compared these results to both prior and concurrent RT-qPCR evaluation (Fig. 2c). By the time of comparative evaluation the number of RT-qPCR positive specimens dropped from 390 to 317, suggesting significant sample degradation either from extended sample storage or multiple freeze-thaw cycles. Nonetheless, mCARMEN was able to identify all 317 (100% sensitivity) of the concurrent RT-qPCR positive specimens. We noted, mCARMEN further detected SARS-CoV-2 in 44 specimens that tested positive by prior RT-qPCR, but were missed by concurrent RT-qPCR testing, suggesting mCARMEN is more robust to sample quality issues. Indeed, if we categorize putative true positives as all specimens that tested positive by prior RT-qPCR as well as present-day RT-qPCR and/or mCARMEN, mCARMEN would have 100% sensitivity compared to 88% for RT-qPCR. mCARMEN also detected SARS-CoV-2 in 3 specimens that tested negative by both prior and concurrent RT-qPCR (Fig. 2e). While we cannot rule out the possibility of false-positives, several pieces of evidence suggest they are more likely to be true positives: mCARMEN demonstrates higher sensitivity over concurrent RT-qPCR testing, these specimens are from suspected SARS-CoV-2 cases based on clinical features, and mCARMEN did not detect SARS-CoV-2 in any clinical specimens prior to the pandemic (Fig. 1, Extended Data Fig. 2).

We further evaluated the analytical sensitivity of RVP by correlating RVP fluorescence signals to Ct values obtained from concurrent RT-qPCR testing (TaqPath, ThermoFisher, CA). Of the 317 specimens positive for SARS-CoV-2 by mCARMEN RVP and both RT-qPCR results, 217 had Ct values <30, suggesting moderate-to-high viral genome copies. By RVP, all 217 specimens (100%) reached signal saturation by 1 hour post-reaction initiation (Fig. 2c&d, Extended Data Fig. 6a&b). The remaining 100 specimens had Ct values between 30-36 and all but 6 samples (94%) reached signal saturation by 1 hour. In total, 98% (311/317) of the specimens reached saturation by 1 hour indicating mCARMEN can rapidly deem viral positivity status for a range of Ct values. Even 17 of the 44 (~40%) RVP positive specimens, but not concurrently RT-qPCR positive, reached saturation by one hour; the slower saturation of these specimens further suggests detection issues caused by sample degradation and/or low viral genome copy number. We also evaluated RVP fluorescence for detecting an internal control and human housekeeping gene, RNase P. We found 525 of the 533 (98.5%) patient specimens reached saturation for RNase P by 1 hour (Extended Data Fig. 6c&d, described in methods).

Additionally, we used unbiased metagenomic NGS as a metric to evaluate RVP performance. As controls for NGS, we sequenced a set of true SARS-CoV-2 negative specimens (i.e., negative by all three results, RVP and 2x RT-qPCR) (n=16), and true SARS-CoV-2 positives (n=15) with a range of Ct values (15-34) (Fig. 2e, Supp. Table 3). Fifteen out of the 16 true negatives had no more than 2 reads mapped to the SARS-CoV-2 genome, in line with <10 reads expected for negative specimens, while 1 specimen had 11 reads by NGS (average ~8.8 million reads per specimen). All true positive specimens had >10 aligned viral reads, ranging from 16 to 802,306 reads, by NGS (100% sensitivity). Only specimens with Ct values <25 (n=8) were able to assemble complete genomes, while specimens with Ct values >25 (n=7) had <200 reads map to SARS-CoV-2.

By NGS, we then evaluated 22 RVP and RT-qPCR discordant specimens, and 8 specimens for which RVP detected other respiratory viruses. The 22 discordant samples included 13 positive by RVP and prior testing but concurrently negative by RT-qPCR, 6 positive by prior testing, but negative by concurrent RVP and RT-qPCR, and 3 positive by RVP but negative by both RT-qPCR results. All but 1 of the 22 (95%) discordant specimens had <10 viral reads by NGS. The single specimen with >10 reads was positive by RVP and prior RT-qPCR, but not concurrent testing, yet just 22 reads mapped to SARS-CoV-2. NGS additionally failed to detect other respiratory viruses in the 8 RVP-positive specimens. RVP identified 4 SARS-CoV-2 co-infections (2 HCoV-HKU1, 1 HPIV-3 and 1 HRSV), and 4 viruses in SARS-CoV-2-negative specimens (3 FLUAV and 1 HCoV-NL63). Given these specimens also had <10 viral reads aligned by NGS, we can neither validate our results as positive nor rule out the possibility of false-negatives by NGS; these samples are likely low viral quantity implying mCARMEN and RT-qPCR are more sensitive.

### Evaluation of RVP performance in a clinical setting

We implemented mCARMEN RVP in the CLIA-certified Clinical Microbiology Laboratory at Massachusetts General Hospital (MGH) to establish assay sensitivity and specificity for clinical validation following FDA guidelines^43^. We first evaluated the limit of detection (LOD), defined as the lowest concentration yielding positive results for at least 19 of 20 replicates. After recapitulating the 9 viral targets on RVP, we found the LOD for HCoV-HKU1, HCoV-NL63, HCoV-OC43, FLUAV, and FLUBV were 500 copies/mL while HMPV and SARS-CoV-2 were 1,000 copies/mL, and HPIV-3 and HRSV were 10,000 copies/mL (Fig. 3a&b, Extended Data Fig. 7a). The LOD likely varies between viral targets for a few reasons: the crRNAs have varying activity levels on their intended target and differing input materials were used based on sample availability.

**Figure 3.**
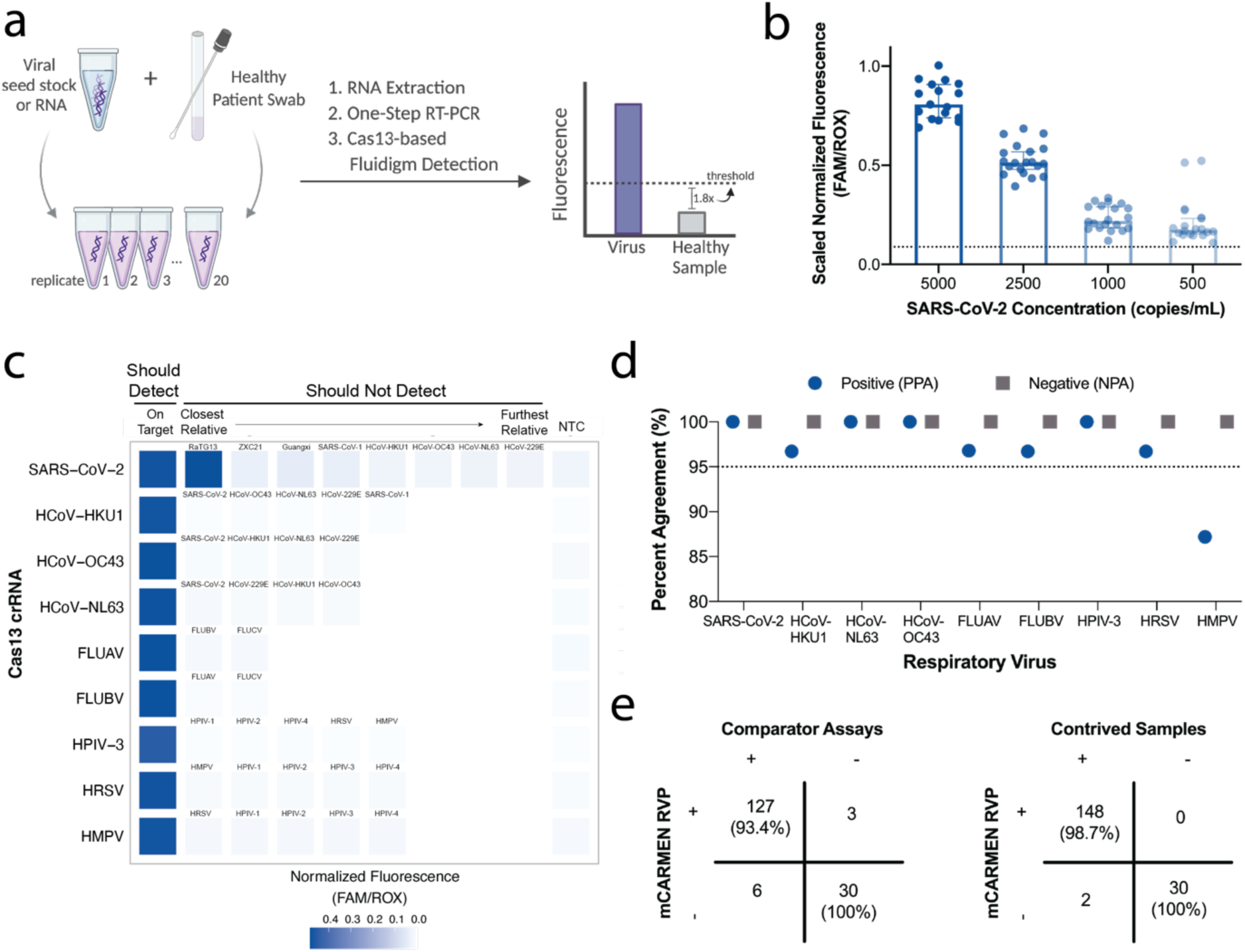
Clinical evaluation of RVP at CLIA-certified laboratory in Massachusetts General Hospital. **a**, Workflow of limit of detection (LOD) studies following the FDA guidelines for establishing assay sensitivity. **b**, Fluorescent values for SARS-CoV-2 target LOD at multiple concentrations, 5,000, 2,500, 1,000, and 500 copies/mL for 20 replicates. **c**, Normalized fluorescence for each virus on RVP evaluated against on-target sequence, closely related sequences (Supp. Table 2), and a no target control, NTC to establish assay specificity. **d**, Positive percent and negative percent agreement (PPA, NPA, respectively) for each virus on RVP calculated based on clinical data in Supp. Table 3. **e**, Concordance of RVP to concurrent comparator assays for the 166 retrospective patient specimens tested (left) and concordance of RVP to contrived samples (right).

After establishing the single analyte LODs, we asked whether co-infections impacted the sensitivity for each virus detected by RVP. To do so, we added SARS-CoV-2 at a constant, 2x LOD, concentration to the remaining 8 viruses on RVP at varying concentrations at or above their respective LOD (Extended Data Fig. 7b). We observed no loss in our ability to detect SARS-CoV-2. However, we noticed a decrease in signal intensity for the other viruses at lower concentrations, yet only one virus, HPIV-3, had a 10-fold higher LOD.

Although we observed no cross-reactivity between RVP panel members in the research setting (Fig. 1 and 2), we followed FDA guidelines to conduct more stringent assay inclusivity and specificity analyses against common respiratory flora and other viral pathogens. *In silico* analysis revealed the primers on RVP are >92% inclusive of the known genetic diversity of each viral species, with additional inclusivity coming from crRNA-target recognition, for an overall >95% inclusivity (Supp. Table 5). When examining off-target activity *in silico*, FDA defines cross-reactivity as >80% homology between one of the primers or probes to any microorganism. We found no more than 75% homology between the RVP primer and crRNA sequences to other closely related human pathogens (Supp. Table 6). This implies that off-target detection will rarely, if ever, occur.

Following *in silico* analysis, we evaluated RVP specificity experimentally. We computationally designed position-matched synthetic gene fragments from closely related viral species, including both human- and non-human-infecting species. When evaluating these gene fragments, only SARS-CoV-2 and RaTG13 showed cross-reactivity (Fig. 3c, Extended Data Fig. 8). This cross-reactivity is expected, however, because the RaTG13 amplicon evaluated shares 100% nucleotide identity with the SARS-CoV-2 amplicon in our assay. We did not observe any cross-reactivity when using viral seed stocks, genomic RNA, or synthetic RNA from ATCC or BEI (Supp. Table 7). Therefore, we found RVP to have 100% analytical specificity.

Finally, the FDA recommends testing a minimum of 30 known-positive clinical specimens for each pathogen in an assay, as well as 30 negative specimens. Where positive specimens are not available, the FDA allows the creation of contrived samples, by spiking viral genomic material at clinically-relevant concentrations into a negative specimen. Each virus evaluated must have a minimum of 95% agreement performance, both positive percent agreement (PPA) and negative (NPA), to clinically-approved comparator assays.

At MGH, archived clinical specimens had been evaluated at the time of collection using one of two comparator assays: Cepheid Xpert Xpress SARS-CoV-2/Flu/RSV multiplexed assay or BioFire RP2.0 multiplexed assay (Extended Data Fig. 9, Supp. Table 3). These included 166 specimens with 137 total viral clinical results: 31 FLUAV, 30 SARS-CoV-2, 30 HRSV, 29 FLUBV, 8 HMPV, 5 HCoV-NL63, 1 FLUBV and HCoV-NL63 co-infection, 1 HCoV-HKU1,1 HCoV-OC43 and 30 clinically negative. Given these specimens can be degraded by multiple freeze thaws, we concurrently tested all specimens by BioFire RP2.0 or TaqPath COVID-19 Combo Kit for the SARS-CoV-2 specimens. We also supplemented this evaluation with 30 contrived samples for each of the following viruses for which we did not have enough positive specimens: HCoV-HKU1, HCoV-OC43, HCoV-NL63, HPIV-3, and HMPV (described in methods), for a total of 150 contrived samples.

All of the RVP viral targets individually had 100% NPA, and all, except HMPV, had >95% PPA to their respective previous comparator assay result, exceeding the minimum clinical performance set by the FDA (Fig. 3d). Of the 137 previously positive clinical results, mCARMEN correctly detected viral nucleic acids 95% (130/137) of the time. For specimens that were evaluated concurrently, mCARMEN and the comparator assay had 9 discordant results (128/137) with equivalent sensitivity for all but the HMPV specimens; BioFire did not detect virus in 3 specimens (1 FLUAV, 1 FLUBV, and 1 HRSV) and mCARMEN did not detect virus in 6 specimens (1 FLUAV, 1 FLUBV, 1 HRSV, and 3 HMPV). Both mCARMEN and BioFire identified 5 specimens with co-infections (HCoV-NL63 in a FLUAV specimen, HPIV-3 in a FLUBV specimen, HCoV-HKU-1 in 2 HRSV specimens, and HCoV-NL63 in a HRSV specimen). Together with the original clinically detected co-infection, there were 6 (1.1%) co-infections in our specimen set (Extended Data Fig. 7c). Overall, mCARMEN and BioFire were 99.4% (1485/1494 individual tests) concordant (Fig. 3e, Extended Data Fig. 9). For the contrived samples, mCARMEN correctly identified 99% (148/150) (Fig. 3e).

We used unbiased metagenomic NGS to further evaluate 9 discordant specimens (2 FLUAV, 2 FLUBV, 2 HRSV, and 3 HMPV), generating an average of 13 million reads per specimen. Either no viral reads were present by NGS or partial genomes were assembled, but the RVP amplicon was missing, making it unlikely for our assay to return a positive result (Extended Data Fig. 9, Supp. Table 3). Based on these results and our previous NGS testing, which indicated NGS was not as sensitive as RVP or the comparator assays, we cannot determine the viral positivity status of these specimens.

### Allelic discrimination distinguishes between SARS-CoV-2 variant lineages of concern

Since current clinical diagnostics are not well positioned to identify mutations - single nucleotide polymorphisms (SNPs), insertions, or deletions - carried in SARS-CoV-2 variant lineages^7,22,23^, we wanted to develop a single platform with both diagnostic and surveillance capabilities for comprehensive detection of 26 SARS-CoV-2 spike gene mutations. We selected these 26 mutations to distinguish between or to detect mutations shared amongst the Alpha, Beta, Gamma, Delta, Epsilon, Lamda, and Omicron variant lineages (Table 1; B.1.1.7, B.1.351, P.1, B.1.617.2, B.1.427/9, C.37, and B.1.1.529 using PANGO nomenclature system, respectively)^8^, and then used a generative sequence design algorithm (manuscript in prep.) to produce crRNAs for allelic discrimination.

**Table 1.** Expected mutations for each SARS-CoV-2 variant lineage.

| Alpha (B.1.1.7) | Beta (1.1.351) | Gamma (P.1) | Delta (B.1.617.2) | Epsilon (B.1.1.427/9) |
| --- | --- | --- | --- | --- |
| Δ69/70 | L18F | L18F | L452R | S13I |
| N501Y | D80A | P26S | T478K | P26S |
| A570D | K417N | K417T | D614G | L452R |
| D614G | E484K | E484K | P681R | D614G |
| P681H | N501Y | N501Y |  |  |
|  | D614G | D614G |  |  |
|  |  | H655Y |  |  |

With the continuous emergence of mutations that can lead to increased transmissibility or enhanced virulence, we also wanted to greatly streamline assay generation for each new SARS-CoV-2 mutation or variant. Thus, we developed an easily adaptable method to track these changes that we call the mCARMEN variant identification panel (VIP). VIP has two non-overlapping primer pair sets within conserved regions of the spike gene to amplify the full-length sequence for use with any crRNA pair. These 26 crRNA pairs, individually or in combination, allow us to track existing variants as well as identify emerging variants, such as Omicron (Fig. 4a). Initially, we tested over 60 combinations of crRNAs on unamplified synthetic material to identify the crRNA pairs with the largest fluorescence ratio for expected divided by unexpected signal at each mutation (Extended Data Fig. 10).

**Figure 4.**
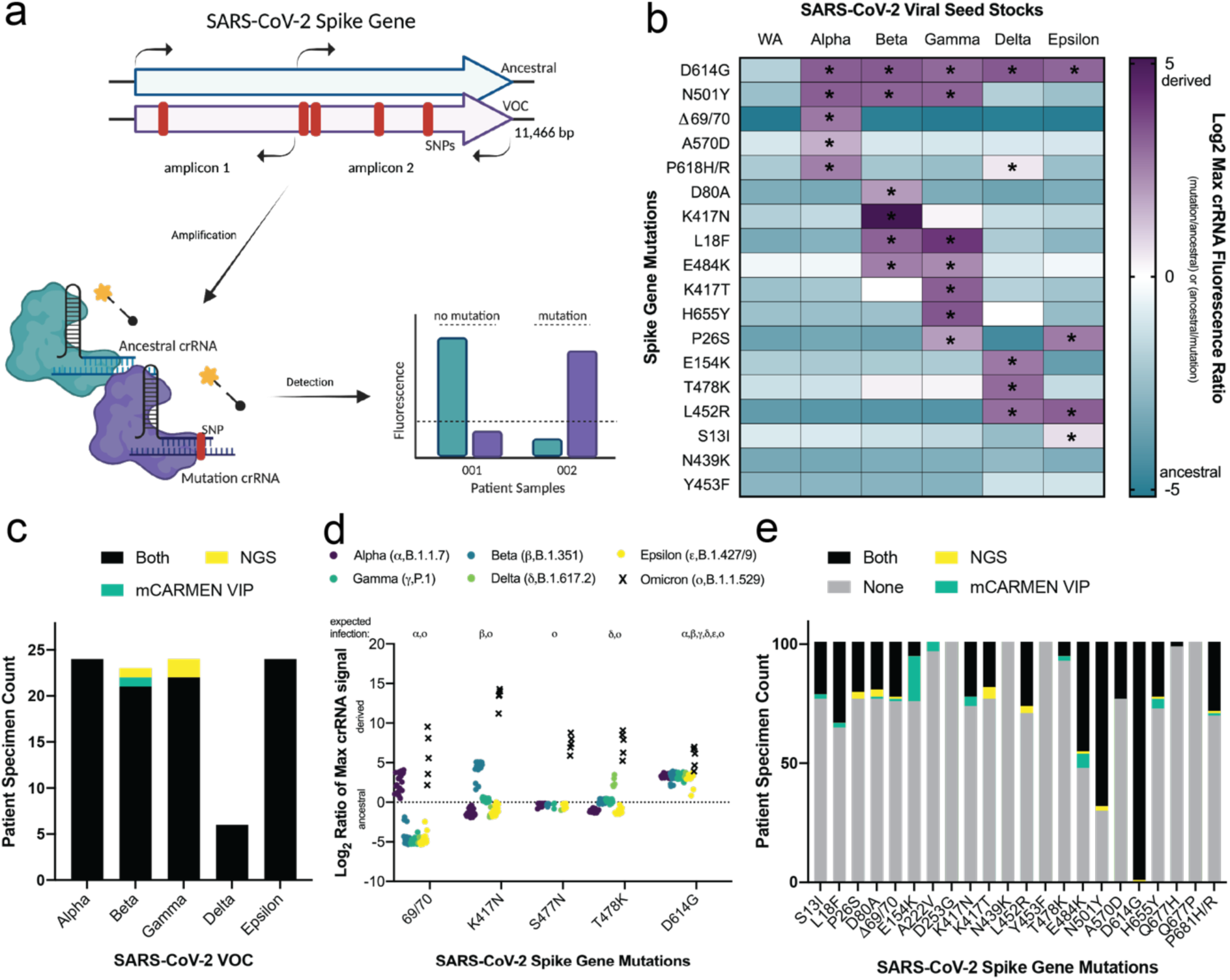
SARS-CoV-2 variant identification using SNP determining Cas13-crRNA combinations. **a**, Schematic of SARS-CoV-2 mutation - SNP or deletion - detection within the spike gene using highly specific Cas13 crRNA designs. **b**, SARS-CoV-2 viral seed stocks from ancestral (WA), Alpha, Beta, Gamma, Delta, or Epsilon variant lineages (10^6^ copies/mL) amplified by 2 primer pairs and tested for the presence or absence of spike gene mutations. Data shown as the log_2_ of the maximum crRNA fluorescence ratio at any time point up to 180 minutes. The ratio that reached maximum fastest is plotted as either ancestral/mutation (blue, negative) or mutation/ancestral (purple, positive). * indicates the particular mutation was confirmed by NGS. **c**, Comparison of 106 SARS-CoV-2 positive variant patient specimens by mCARMEN and NGS based on the final variant call assessed by unique combinations of mutations (Methods Variant Calling). Black: both VIP and NGS; Yellow: NGS only; Green: VIP only. **d**, 106 variant patient specimens tested for various SNPs by mCARMEN are organized by Alpha, Beta, Gamma, Delta, or Epsilon. Data shown as the log_2_ maximum crRNA fluorescence ratio of mutation/ancestral (positive) or ancestral/mutation (negative) at any time point up to 180 minutes focusing on the Omicron mutations. Ancestral <0, derived >0, dashed line at 0 for reference. **e**, Individual mutation breakdown of mCARMEN results to NGS on 106 variant patient specimens. Black: mutations shared by both VIP and NGS; Yellow: NGS only; Green: VIP only; Gray: ancestral for VIP and NGS.

We validated the flexible VIP method by testing RNA extracted from SARS-CoV-2 viral seed stocks, for the ancestral (Washington isolate: USA-WA1, ATCC) lineage, and Alpha, Beta, Gamma, Delta, and Epsilon lineages (Fig. 4b, Extended Data Fig. 11). As expected, the WA SARS-CoV-2 viral seed stock isolate showed ancestral signals for all mutations tested. Alpha, Beta, Gamma, Delta, and Epsilon had expected signals for every mutation confirmed by NGS (Table 1, description in methods). Though each crRNA has different kinetics owing to varying hit-calling thresholds, we almost always observed a higher expected signal above the unexpected signal, which is important in the prevention of false positive results (Extended Data Fig. 12).

For clinical relevance, we developed an automated variant calling procedure that evaluates the mutation-specific signal in SARS-CoV-2-positive patient specimens and returns a variant lineage result (Extended Data Fig. 13a, described in methods). For some mutations at the same or similar genomic position we observed cross-reactive signals which we overcame by comparing the maximum fluorescent ratios between those mutations and assigning the positive call to the higher of the two (Extended Data Fig. 13b).

We applied VIP and the analysis pipeline to identify the variant lineage in 106 known SARS-CoV-2-positive patient specimens: 24 Alpha, 23 Beta, 24 Gamma, 6 Delta, and 24 Epsilon, and 5 presumptive Omicron specimens. Of the 101 specimens with NGS results, excluding Omicron specimens, all but 3 (97%) specimens (1 Beta and 2 Gammas) were given the correct variant lineage identification (Fig. 4c&d, Extended Data Fig. 13c, Supp. Table 3). The Beta specimen had signal for a Beta-specific SNP, K417N, but also had signal for E154K, a Delta-specific SNP. The Gamma specimens had no unique signals and shared signals for mutations overlapping with the Beta lineage resulting in a “Variant not Identified” call. The 5 presumed Omicron specimens had S gene drop-out by TaqPath RT-qPCR and were found to have the following mutations by mCARMEN: del69/70, K417N, S477N, T478K, N501Y, and D614G (Fig. 4d). mCARMEN is able to return a result within a few hours while NGS takes several days thus we are still awaiting the results for final comparison.

Focusing on the results for the individual mutations themselves, we found that only 2 mutations, E154K and E484K, had more than 5 specimens differ in their results between NGS and VIP (Fig. 4e). E154K had 19 differences, all for Epsilon specimens, while E484K had 7 differences. Since all Epsilon specimens contain a SNP 2 amino acids away from E154, we believe the E154K crRNA is detecting the Epsilon W152C mutation, which would account for all 19 discrepancies between NGS and VIP. Meanwhile the 7 E484K discrepancies are likely attributed to our comparison of cross-reactive signals between E484K and T478K, thus new crRNA designs are likely needed to optimally differentiate these signals. Altogether, we found VIP had 97.9% concordance to NGS at allelic discrimination.

### Quantification of viral genomic copies in samples using Cas12 and Cas13 reaction kinetics

Similar to widely-used multiplexed approaches, like BioFire^17^, the original design of CARMEN^37^ did not provide a true quantitative assessment of viral genome copies present in a sample. Establishing the total viral quantity in a patient is important for assessing the stage of infection, transmission risk, and most effective treatment plan^9,10^. The gold standard assay for sample quantification, RT-qPCR, leverages the standard curve - serial dilutions of a given target at a known concentration - as a means of using Ct values to approximate viral quantity^44^. We wanted to determine if a similar approach could be applied to mCARMEN.

To make mCARMEN quantitative, we took advantage of the existence of multiple CRISPR/Cas proteins with differing reaction kinetics and enzymatic activities, and the 3 fluorescent channels detected by the Fluidigm Biomark (Fig. 5a). We incorporated DNA-targeting CRISPR/Cas12 into the Cas13 reaction, and used protein-specific reporters in different fluorescent channels, HEX and FAM, respectively to maximize our multiplexing capabilities. To capture reaction kinetics, images of the IFC chip are taken every 5 minutes for 3 hours to generate sigmoidal curves from the fluorescent signals over time. When considering enzymatic activities, Cas13 has enhanced sensitivity compared to Cas12 since the process of reverse transcribing the dsDNA sample input for Cas13 detection results in increased starting concentration. Thus, we use Cas12 to capture the kinetic curves of higher copy material on the standard curve and Cas13 to capture lower copy material.

**Figure 5.**
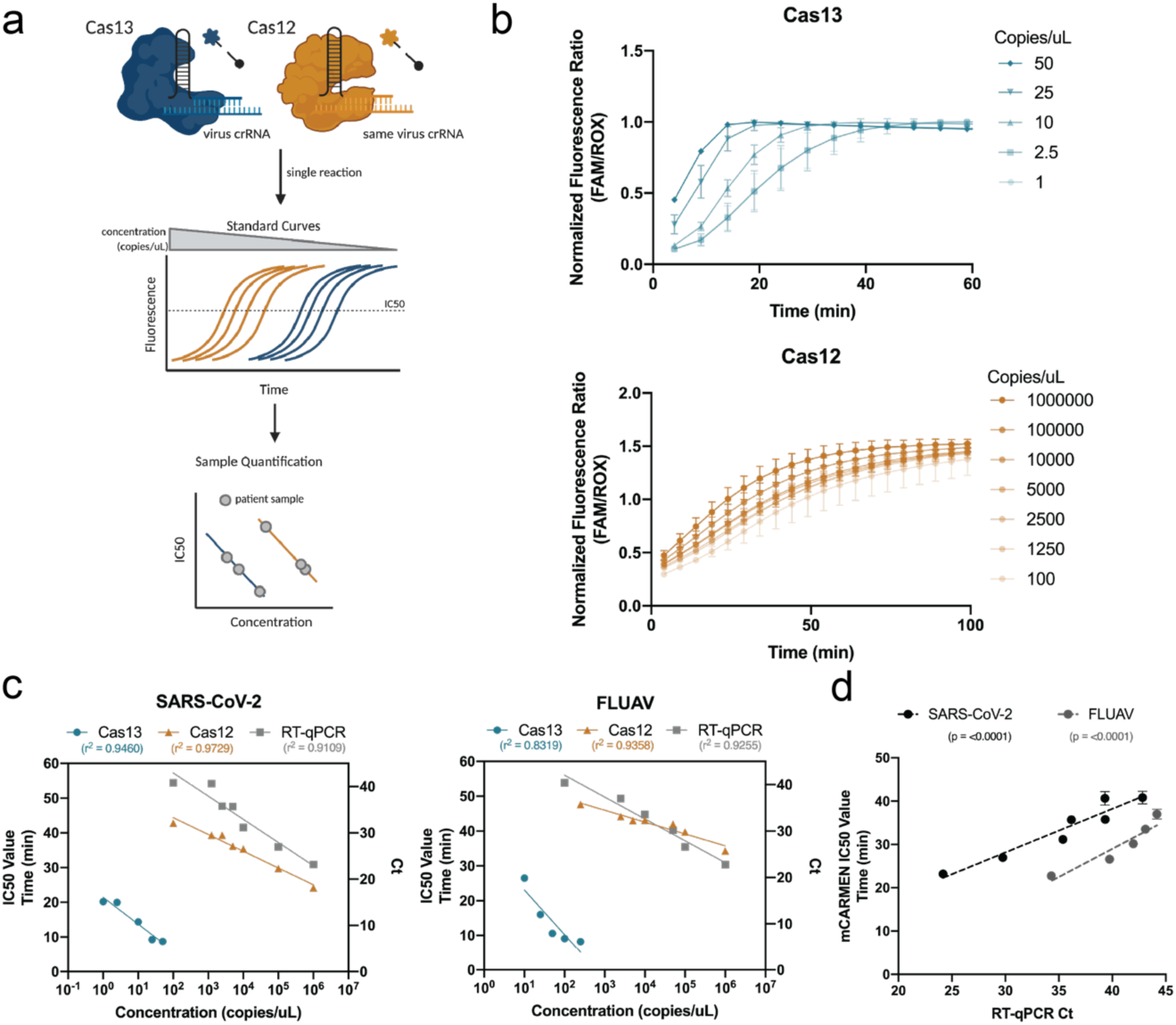
Viral quantification in samples with RVP using both Cas12 and Cas13 in combination. **a**, Schematic of Cas12 and Cas13 dual detection on mCARMEN to quantify viral copy number in samples. Similar to RT-qPCR, standard curves are utilized for assay quantification. Cas12 captures higher copy material while Cas13 captures lower copy material. Fluorescence is plotted over time to determine the IC_50_ value at each concentration using a sigmoidal 4PL fit. The IC_50_ values are then plotted by concentration to generate a semilog line with an R^2^ value >0.8 for Cas12 and Cas13 individually. After line generation, the IC_50_ value of each patient sample is plotted onto these lines to determine viral copies/μL. **b**, Normalized fluorescence ratio of Cas13-crRNA (top) or Cas12-crRNA (bottom) signal over time at varying concentrations of synthetic SARS-CoV-2 Orf1ab RNA. **c**, Semilog lines generated by IC_50_ values from Cas12 and Cas13 crRNA signals, and Ct values from RT-qPCR for >4 target concentrations of SARS-CoV-2 and FLUAV IVT RNA. Blue: Cas13; Orange: Cas12; Gray: RT-qPCR. **d**, Linear regression to compare mCARMEN IC_50_ values to RT-qPCR Ct values with a best line fit shown as a dashed line. Black: SARS-CoV-2; Gray: FLUAV.

We integrated our quantification efforts into RVP, since this assay was extensively evaluated in both research and clinical settings. We manually designed Cas12 crRNAs in the same region of the viral genome that the RVP Cas13 crRNAs target for a two-step standard curve generation on the same target amplicon for Cas12- and Cas13- RNPs individually. The first step requires plotting the fluorescence for a range of concentrations (Cas12: 10^7^-10^3^ copies/μL; Cas13: 10^3^-10^0^ copies/μL) at each time point to calculate the IC_50_ through a sigmoidal, four parameter logistic (4PL) curve, R^2^ >0.9 (Fig. 5b, Extended Data Fig. 14a&b). In some cases, we could not determine the IC_50_ value because signal saturation occurred too quickly or not at all and therefore, that concentration was excluded from analysis. In the second step, we plotted the IC_50_ values onto a semilog line, where concentration is logarithmic and time is linear, to generate the standard curves (Fig. 5c). We compared these results to a standard curve generated from RT-qPCR using the same serial dilutions and found a linear relationship between SARS-CoV-2 and FLUAV IC_50_ values to Ct values (R^2^ 0.901 and 0.881, respectively) (Fig. 5d). In all, these results suggest that by using Cas12 and Cas13 in combination, we could extrapolate viral quantification - spanning a 10^0^-10^6^ range of target concentrations - from patient specimens with performance similar to RT-qPCR.

## Discussion

Here, we report mCARMEN, a high-throughput, multiplexed, and microfluidic diagnostic and surveillance platform with panels for respiratory viruses and SARS-CoV-2 variants that can be parallelized to test 300-550 patient specimens in an 8 hour working day. To make mCARMEN a clinically relevant technology, we built on CARMEN v1^37^ by streamlining the workflow and incorporating commercially available Fluidigm instrumentation^38^. We validated mCARMEN on 902 patient specimens for the detection of 9-21 human respiratory viruses (RVP) or SARS-CoV-2 variant mutations (VIP) with high concordance to comparator assays which passed the FDA’s performance criteria^43^ for all but one virus. Notably, when testing previously positive clinical specimens, we found a substantial proportion were not positive by concurrent testing, but were positive by mCARMEN (Fig. 2). This suggests sample degradation issues - a known problem when detecting RNA viruses in clinical specimens^41,45^ - that mCARMEN is more robust to handling than RT-qPCR or NGS. Though we cannot rule out false positives, we did not detect SARS-CoV-2 in specimens prior to the pandemic and we had 100% concordance with true virus-negative specimens.

To enhance mCARMEN’s clinical diagnostic relevance and meld it with surveillance technology requirements, we further maximized its multiplexing capabilities by discriminating between mutations for variant lineage classification in patient specimens and quantifying viral genomic copies. Currently, variant lineage classification is only evaluated by NGS, which is costly and relies on specialized expertise found outside the clinic^22,24^. VIP gives similarly rich information about key SARS-CoV-2 mutations at 5-10x cheaper, per sample, than NGS, and is far more comprehensive than current nucleic acid-based diagnostics. Importantly, VIP was poised to differentiate Omicron immediately as it emerged since we routinely design guides to preemptively identify mutations of interest in the spike gene in preparation for emerging variants. Given the number of mutations detected by VIP, we expect to observe distinct mutation signatures between variant lineages that will allow us to differentiate these and future variants of concern from each other without assay redesign. We also adapted mCARMEN for dual Cas12 and Cas13 detection by capitalizing on the differing protein kinetics. A few groups have studied Cas12 and Cas13 reaction kinetics to inform assay quantification^46,47^, but the range of concentrations being quantified has been limited due to reaction saturation. We expanded the quantifiable concentration range to 5-6 orders of magnitude, which is similar to RT-qPCR. These mCARMEN applications have the potential to provide a more holistic diagnosis to the patient.

We rapidly developed mCARMEN for use in the pandemic, but faced challenges during the clinical validation and approval process needed for a large-scale roll-out. Specifically, it was difficult to obtain at least 30 previously confirmed clinical specimens for each virus on RVP with enough material available for extensive concurrent testing, while also facing specimen degradation issues that inevitably occur over time. mCARMEN’s performance exceeded FDA requirements, yet the FDA was unable to give it full emergency use authorization (EUA) review and approval because of their limited bandwidth and need to prioritize other applications. Lastly, funding to support mCARMEN in clinics and beyond has been scarce despite there being a clear need for it.

Although further work would be required to bring mCARMEN fully to the clinic, mCARMEN can function as an ideal single technology platform with diagnostic and surveillance capabilities for detecting respiratory pathogens and variants. We have taken significant steps to streamline assay workflow while enhancing sensitivity and not sacrificing specificity. There is currently no other diagnostic technology that combines multiplexed pathogen testing with variant tracking and is highly scalable and amenable to clinical laboratory settings. This technology has the potential to test for other infectious diseases^48^ and sample types for an even more comprehensive, multiplexed, and high-throughput diagnostic, all while maintaining a level of clinical relevance that is unmatched to other nucleic acid- or antigen-based diagnostics.

## Data Availability

All requests for raw and analyzed data and materials will be reviewed by the Broad Institute of Harvard and MIT to verify if the request is subject to any intellectual property or confidentiality obligations. Data and materials that can be shared will be released via a Material Transfer Agreement. RNA sequencing data have been deposited to the Sequence Read Archive under the accession code (to be updated) and will be made available upon request for academic use and within the limitations of the provided informed consent by the corresponding author upon acceptance. Source code is available on github: https://github.com/broadinstitute/mcarmen.

## Author Contributions

N.L.W. and C.M. initially conceived this study then involved C.M.A., S.G.T., and J.W. for preliminary implementation. N.L.W., M.Z., J.W., C.M.A., S.G.T., M.W.T., J.K. set up 21 respiratory virus testing on CARMEN v1. N.L.W. and S.M. designed the primers and crRNAs for the 21 respiratory viruses tested on CARMEN v1 and mCARMEN. N.L.W., J.W., C.M.A., S.G.T. performed initial experiments on Fluidigm instrumentation. M.Z., J.W., C.M.A. wrote the python scripts for mCARMEN data analysis. N.L.W. performed experiments to streamline the mCARMEN workflow with help from C.H., J.W., and S.G.T. N.L.W., C.H., MM conducted the SARS-CoV-2 patient sample testing in an academic setting, B.L.M. and F.C. helped obtain these samples. C.H., E.M.M., B.M.S., J.E., D.B., G.M. performed clinical evaluation of mCARMEN RVP at MGH, under guidance from N.L.W., J.J., J.E.L., E.R., J.A.B., C.M. J.W. wrote and generated the software used for RVP. N.L.W. with help from M.R.B. conducted NGS on patient samples. N.L.W. designed and tested the primers for VIP, and N.L.W. and S.M. designed the crRNAs for VIP. N.L.W. conducted the experimental validation of VIP. M.K.K., M.W.W., M.W.K., and J.R.B. provided FLUAV samples, SARS-CoV-2 VOC seed stocks, and SARS-CoV-2 VOC patient samples and the corresponding NGS data. K.J.S. provided assistance in patient specimen collection. M.S., S.J., M.L.C. provided Omicron specimens. N.L.W. tested all SARS-CoV-2 VOC specimens by mCARMEN. M.Z. wrote and generated the VOC calling analysis pipeline for VIP testing under guidance from N.L.W. N.L.W. conducted experiments to make mCARMEN quantitative with assistance from T.G.N. K.J.S., B.L.M., C.T.H., D.T.H., J.E.L., J.R.B., E.R., J.A.B., P.C.B., P.C.S., C.M. provided insights into the work overall. N.L.W. generated the figures with help from M.Z. and J.W. N.L.W. wrote the paper with help from C.H. and guidance from P.C.S. and C.M. J.A.B., P.C.B., P.C.S., and C.M. jointly supervised the work. All authors reviewed the manuscript.

## Acknowledgements

We would like to thank the Blainey Lab and Hung Lab at the Broad Institute for providing additional laboratory space to perform the work; M. Kirby, M. Wilson, M. Keller, J. Barnes for providing FLUAV samples, SARS-CoV-2 variant seed stocks, and SARS-CoV-2 variant patient samples and the corresponding NGS data. J. Arizti Sanz, Y. Zhang, and A. Bradley for helping with guide design or sharing reagents; G. Adams, S. Dobbins, K. DeRuff and other members of the Sabeti Lab COVID-19 sequencing team for providing patient samples; L. Krasilnikova, C. Tomkins-Tinch, C. Loreth, and other Sabeti Lab members for providing assistance with NGS analysis on Terra; B. Zhou for helping provide the SARS-CoV-2 VOC seed stocks and patient samples; H. Metsky, C. Freije, J. Arizti Sanz, S. Siddiqui for their thoughtful discussions and reading of the manuscript. Funding was provided by DARPA D18AC00006. This work is made possible by support from Flu lab and a cohort of generous donors through TED’s Audacious Project, including the ELMA Foundation, MacKenzie Scott, the Skoll Foundation, and Open Philanthropy. Funding for NGS was provided by Centers for Disease Control and Prevention COVID-19 baseline genomic surveillance contract sequencing (75D30121C10501 to Clinical Research Sequencing Platform, LLC), a CDC Broad Agency Announcement (75D30120C09605 to B.L.M), National Institute of Allergy and Infectious Diseases (U19AI110818 to P.C.S). C.M. is supported by start-up funds from Princeton University. M.Z. and M.W.T. were supported by the National Science Foundation Graduate Research Fellowship under Grant No. 1745302. B.A.P. is supported by the National Institute of General Medical Sciences grant T32GM007753.

The views, opinions, conclusions, and/or findings expressed should not be interpreted as representing the official views or policies, either expressed or implied, of the Department of Defense, US government, National Institute of General Medical Sciences, DHS, or the National Institutes of Health. The DHS does not endorse any products or commercial services mentioned in this presentation. In no event shall the DHS, BNBI or NBACC have any responsibility or liability for any use, misuse, inability to use, or reliance upon the information contained herein. In addition, no warranty of fitness for a particular purpose, merchantability, accuracy or adequacy is provided regarding the contents of this document. The United States Government retains and the publisher, by accepting the article for publication, acknowledges that the United States Government retains a non-exclusive, paid up, irrevocable, world-wide license to publish or reproduce the published form of this manuscript, or allow others to do so, for United States Government purposes.

The findings and conclusions in this report are those of the authors and do not necessarily represent the official position of the Centers of Disease Control and Prevention (CDC). Use of trade names and commercial sources is for identification purposes only and does not imply endorsement.

## Competing Interests Statement

N.L.W., S.G.T., C.M.A., D.T.H., P.C.B., P.C.S., and C.M. are co-inventors on a patent related to this work. P.C.B is a co-inventor on patent applications concerning droplet array technologies and serves as a consultant and equity holder of companies in the microfluidics and life sciences industries, including 10x Genomics, GALT, Celsius Therapeutics, Next Generation Diagnostics, Cache DNA, and Concerto Biosciences; P.C.B’s laboratory receives funding from industry for unrelated work. P.C.S. is a co-founder of and consultant to Sherlock Biosciences and a Board Member of Danaher Corporation, and holds equity in the companies.

## Code availability

The code used for data analysis in this study is made available on Github: https://github.com/broadinstitute/mcarmen.

## Data availability

All requests for raw and analyzed data and materials will be reviewed by the Broad Institute of Harvard and MIT to verify if the request is subject to any intellectual property or confidentiality obligations. Data and materials that can be shared will be released via a Material Transfer Agreement. RNA sequencing data will be made available upon request for academic use and within the limitations of the provided informed consent by the corresponding author upon acceptance. Source code is available on github: https://github.com/broadinstitute/mcarmen.

## Reporting summary

Further information on research design is available in the Nature Research Reporting Summary linking to this article.

## Methods

### 1. Patient samples / ethics statement

Use of clinical excess of human specimens from patients with SARS-CoV-2 from the Broad Institute’s Genomics Platform CLIA Laboratory was approved by the MIT IRB Protocol #1612793224. Additional SARS-CoV-2 samples were collected from consented individuals under Harvard Longwood Campus IRB #20-1877 and covered by an exempt determination (EX-7295) at the Broad Institute. Other human-derived samples from patients with SARS-CoV-2 were collected by the CDC and determined to be non-human subjects research; the Broad Office of Research Subject Protections determined these samples to be exempt. Human specimens from patients with SARS-CoV-2, HCoV-HKU1, HCoV-NL63, FLUAV, FLUBV, HRSV, and HMPV were obtained under a waiver of consent from the Mass General Brigham IRB Protocol #2019P003305. Researchers at Princeton were determined to be conducting not-engaged human subjects research by the Princeton University IRB.

### 2. General mCARMEN Procedures

#### 2a. Preparation and handling of synthetic materials

crRNAs were synthesized by Integrated DNA Technologies (Coralville, IA), resuspended in nuclease-free water to 100 μM, and further diluted for input into the detection reaction. Primer sequences were ordered from Eton or Integrated DNA Technologies, resuspended in nuclease-free water to 100 μM, and further combined at varying concentrations for pooled amplification.

#### 2b. Preparation of *in vitro* transcribed (IVT) material

DNA targets were ordered from Integrated DNA Technologies and *in vitro* transcribed (IVT) using the HiScribe T7 High Yield RNA Synthesis Kit (New England Biolabs, NEB). Transcriptions were performed according to the manufacturer’s recommendations with a reaction volume of 20 μL that was incubated overnight at 37°C. The transcribed RNA products were purified using RNAClean XP beads (Beckman Coulter) and quantified using NanoDrop One (Thermo Scientific). Depending on the experiment, the RNA was serially diluted from 10^11^ down to 10^-3^ copies/μL and used as input into the amplification reaction.

#### 2c. Extraction - Manual or Automated

RNA was manually extracted from input material using the QIAamp Viral RNA Mini Kit (QIAGEN) according to the manufacturer’s instructions. RNA was extracted from 140 μl of input material with carrier RNA and samples were eluted in 60 μl of nuclease free water and stored at −80 °C until use.RNA was automatically extracted using the MagMAX™ DNA Multi-Sample Ultra 2.0 Kit on a KingFisher™ Flex Magnetic Particle Processor with 96 Deep-Well Head (Thermo Fisher Scientific). RNA was extracted from 200 µL of input material and was run according to the “Extract RNA - Automated method (200-μL sample input volume)” protocol in <u>TaqPath™ COVID-19 Combo Kit Protocol</u>, on page 21-24. The MVP_2Wash_200_Flex protocol was used. Samples were eluted in 50 μl of elution solution and either directly added to the amplification reaction or stored at −80 °C until use.

#### 2d. Amplification: Qiagen or SSIV

We followed the CARMEN v1 platform for two-step reverse transcription amplification then transitioned to a single-step amplification reaction after experiments depicted in Figure 1. We used the Qiagen OneStep RT-PCR Mix for Figures 2,3,5 and Invitrogen SuperScript IV One-Step RT-PCR System for Figure 4. For the Qiagen OneStep RT-PCR, a total reaction volume of 50 μL was used with some modifications to the manufacturers recommended reagent volumes. Specifically 1.25X final concentration of OneStep RT-PCR Buffer, 2x more Qiagen Enzyme Mix, and 20% RNA input. Final concentrations for all viral primers were 300 nM and 100 nM for RNase P primers. The following thermal cycling conditions were used: (1) reverse transcription at 50°C for 30 min; (2) initial PCR activation at 95°C for 15 min; (3) 40 cycles of 94°C for 30 s, 58°C for 30 s, and 72 °C for 30 s. For Invitrogen SuperScript IV One-Step RT-PCR, a total reaction volume of 25 μL with 20% RNA input and final primer concentrations at 1 μM. The following thermal cycling conditions were used: (1) reverse transcription at 50°C for 10 min; (2) initial PCR activation at 98°C for 2 min; (3) 35 cycles of 98°C for 10 s, 60°C for 10 s, and 72 °C for 1 min 30 s; (4) final extension at 72 °C for 5 min. See Supp. Table 2 for information on primer sequences used in each mCARMEN panel.

#### 2e. Fluidigm Detection

The Cas13 detection reactions were made into two separate mixes: assay mix and sample mix, for loading onto a microfluidic IFC (depending on the experiment, either Gene Expression (GE) or Genotyping (GT) IFCs were used in either a 96.96 or 192.24 format) (Fluidigm):

The *assay mix* contained LwaCas13a (GenScript) and on occasion LbaCas12a (NEB) concentration varied with experiment, 1× Assay Loading Reagent (Fluidigm), 69U T7 polymerase mix (Lucigen), and crRNA concentration varied with experiment for a total volume of 16 μL per reaction. See below for details pertaining to each mCARMEN panel.

The *sample mix* contained 25.2U RNase Inhibitor (NEB), 1× ROX Reference Dye (Invitrogen), 1× GE Sample Loading Reagent (Fluidigm), 1 mM ATP, 1 mM GTP, 1 mM UTP, 1 mM CTP, 9 mM MgCl_2_ in a nuclease assay buffer (40 mM Tris-HCl pH 7.5, 1 mM DTT) and either a 500 nM quenched synthetic fluorescent RNA reporter (FAM/rUrUrUrUrUrUrU/3IABkFQ/ or VIC/rTrTrArTrTrArTrT/3IABkFQ/ Integrated DNA Technologies) or RNaseAlert v2 (Invitrogen) was used for a total volume of 12.6 μL. See below for details on each mCARMEN panel.

##### IFC Loading and Run

Syringe, Actuation Fluid, Pressure Fluid (Fluidigm), and 4 μL of assay or sample mixtures were loaded into their respective locations on a microfluidic IFC (depending on the experiment, either Gene Expression (GE) or Genotyping (GT) IFCs were used in either a 96.96 or 192.24 format) and were run according to the manufacturer’s instructions. The IFC was loaded onto the IFC Controller RX or Juno (Fluidigm) where the ‘Load Mix’ script was run. After proper IFC loading, images over either a 1-3 hour period were collected using a custom protocol on Fluidigm’s EP1 or Biomark HD.

#### 2h. Fluidigm Data Analysis

We plotted reference-normalized background-subtracted fluorescence for guide-target pairs. For a guide-target pair (at a given time point, t, and target concentration), we first computed the reference-normalized value as (median(P_t_-P_0_)/(R_t_-R_0_)) where P_t_ is the guide signal (FAM) at the time point, P_0_ is its background measurement before the reaction, R_t_ is the reference signal (ROX) at the time point, R_0_ is its background measurement, and the median is taken across replicates. We performed the same calculation for the no template (NTC) control of the guide, providing a background fluorescence value for the guide at t (when there were multiple technical replicates of such controls, we took the mean value across them). The reference-normalized background-subtracted fluorescence for a guide-target pair is the difference between these two values.

### 3. 21 Respiratory Viruses (Fig. 1)

#### 3ai. Design

The oligonucleotide primers and CRISPR RNA guides (crRNAs) are designed for detection of conserved regions of the following respiratory viruses: SARS-CoV-2, HCoV-229E, HCoV-HKU1, HCoV-NL63, HCoV-OC43, FLUAV, FLUBV, HMPV, HRSV, HPIV-1,2,3,4, AdV, HEV-A,B,C,D, SARS-CoV, MERS-CoV, and HRV. More specifically, complete genomes for all viruses on the panel were downloaded from NCBI and aligned using MAFFT ^49^. For viral species with fewer than 1000 sequences, MAFFT’s ‘FFT-NS-ix1000’ algorithm was used. For viral species with >1000 sequences, MAFFT’s ‘FFT-NS-1’ algorithm was used. These aligned sequences were then fed into ADAPT for crRNA design with high coverage using the ‘minimize guides’ objective (>90% of sequences detected). Once highly conserved regions of the viral genome were selected with ADAPT for optimal guide design, primers were manually designed to amplify a 100-250 bp target region with the crRNA predicted to bind in the middle of the fragment. ADAPT’s constraints on primer specificity were relaxed and in some cases multiple primers were needed to encompass the full genomic diversity of a particular virus species. For optimal amplification, the primers were split into two pools. These primer pools and crRNA sequences are listed in Supp. Table 2.

#### 3aii. Target control - PIC1 and PIC2

The consensus sequences generated directly above after multiple genome alignment with MAFFT were used to order a 500 bp dsDNA fragment encompassing the primer and crRNA binding sites. RNA was generated following the method described in ‘General mCARMEN Procedure - Preparation of IVT Material’ and diluted to 10^6^ copies/μl in pools based on the primer pools mentioned above (PIC1, PIC2). The PICs were used as input into the CARMEN v1 or mCARMEN detection reaction to function as a detection positive control.

#### 3b. Sample Extraction: Manual or Automated

Automated and manual extraction was performed according to methods described under ‘General mCARMEN Procedures - Extraction’

#### 3c. Amplification: Two-Step

We followed the CARMEN v1 platform for two-step reverse transcription amplification, which was performed first by cDNA synthesis and then by PCR.

##### 3ci. cDNA synthesis using SSIV

10 μL of extracted RNA was converted into single-stranded cDNA in a 40 μL reaction. First, Random Hexamer Primers (ThermoFisher) were annealed to sample RNA at 70 °C for 7 min, followed by reverse transcription using SuperScript IV (Invitrogen) for 20 min at 55 °C. cDNA was stored at −20 °C until use. DNase treatment was not performed at any point during sample preparation.

##### 3cii. Q5 DNA amplification

Nucleic acid amplification was performed via PCR using Q5 Hot Start polymerase (NEB) using primer pools (with 150 nM of each primer) in 20 μL reactions. Amplified samples were added directly into the detection reaction or stored at −20 °C until use. The following thermal cycling conditions were used: (1) initial denaturation at 98 °C for 2 min; (2) 45 cycles of 98 °C for 15 s, 50 °C for 30 s, and 72 °C for 30 s; (3) final extension at 72 °C for 2 min. Each target was amplified with its corresponding primer pool, as listed under ‘oligonucleotides used in this study.’

#### 3d. Detection

##### 3di. CARMEN-Droplet

For colour coding, unless specified otherwise, amplified samples were diluted 1:10 into nuclease-free water supplemented with 13.2 mM MgCl_2_ prior to colour coding to achieve a final concentration of 6 mM after droplet merging. Detection mixes were not diluted. Colour code stocks (2 µl) were arrayed in 96W plates (for detailed information on construction of colour codes, see ‘Colour code design, construction and characterization’). Each amplified sample or detection mix (18 µl) was added to a distinct colour code and mixed by pipetting.

For emulsification, the colour-coded reagents (20 µl) and 2% 008-fluorosurfactant (RAN Biotechnologies) in fluorous oil (3M 7500, 70 µl) were added to a droplet generator cartridge (Bio Rad), and reagents were emulsified into droplets using a Bio Rad QX200 droplet generator or a custom aluminum pressure manifold.

For droplet pooling, a total droplet pool volume of 150 µl of droplets was used to load each standard chip; a total of 800 µl of droplets was used to load each mChip. To maximize the probability of forming productive droplet pairings (amplified sample droplet + detection reagent droplet), half the total droplet pool volume was devoted to target droplets and half to detection reagent droplets. For pooling, individual droplet mixes were arrayed in 96W plates. A multichannel pipette was used to transfer the requisite volumes of each droplet type into a single row of eight droplet pools, which were further combined to make a single droplet pool. The final droplet pool was pipetted up and down gently to fully randomize the arrangement of the droplets in the pool. The pooling step is rapid (<10 min), and small molecule exchange between droplets during this period does not substantially alter the colour codes.

##### 3dii. mCARMEN

We followed the methods under General CARMEN Procedures - Detection - Fluidigm detection with the following modifications: 42.5 nM LwaCas13 and 212.5 nM crRNA in each assay mix reaction, and 500 nM RNaseAlert v2 in each sample mix reaction.

#### 3e. Data Analysis

##### 3ei. CARMEN v1

We followed the data analysis pipeline from CARMEN v1 to demultiplex and readout the fluorescence intensity of the reporter channel for each droplet reaction performed (REF CARMEN v1). In brief, pre-merge imaging data was processed using custom Python scripts to detect fluorescently-encoded droplets in microwells and identify their inputs based on their fluorescence intensity in three encoding channels, 647 nm, 594 nm, and 555 nm. Subsequently, post-merge imaging data was analyzed to extract the reporter signal of the assay in the 488 nm channel, and those reporter fluorescence intensities were physically mapped to the contents of each microwell. Quality control filtering was performed based on the appropriate size of a merged droplet from two input droplets and the closeness of a droplet’s color code to its assigned color code cluster centroid. The median and standard error were extracted from the replicates of all assay combinations generated on the array.

##### 3eii. mCARMEN

We followed the methods under ‘General CARMEN Procedures - Fluidigm Data Analysis’ and further visualized the data using python, R, and Prism.

### 4. Single-step amplification troubleshooting

The following RT-PCR kits were tested to determine the best performing assay: (1) OneStep RT-PCR Kit (Qiagen) (2) TaqPath™ 1-Step Multiplex Master Mix (Thermo Fisher), (3) One Step PrimeScript™ RT-PCR Kit (Takara), (4) GoTaq® Probe RT-qPCR Kit (Promega), (5) UltraPlex™ 1-Step ToughMix® (4X) (Quantbio), (6) iTaq™ Universal One-Step Kits for RT-PCR (Bio-Rad). Of the kits tested, the OneStep RT-PCR Kit (Qiagen) was chosen for the final mCARMEN protocol.

#### 4a. OneStep RT-PCR Kit (Qiagen)

All or a combination of the following thermal cycling condition ranges were tested to shorten assay run-time: reverse-transcription at 50 °C at 15-30 min, PCR activation at 95 °C at 5-15 min, denaturation step at 94 °C at 10-30 s, and extension step at 72 °C for 10 s to 1 min. The final extension at 72 °C for 10 min was omitted in all runs. The following primer pool conditions were also tested to optimize the assay: 150 nM, 300 nM, 500 nM, and 600 nM of virus-specific primer and 100 nM and 150 nM of RNaseP primers, with 5 μM of each virus-specific primer and 1.7 μM of RNaseP primers. Reaction volumes tested include: 20 μl with 10% RNA template input, 30 μl with 20% RNA template input, and 50 μl with 20% RNA template input. The final amplification conditions used for the RVP panel are described under ‘General mCARMEN Procedures - Amplification’.

#### 4b. TaqPath™ 1-Step Multiplex Master Mix Kit (Thermo Fisher)

The **TaqPath™** 1-Step Multiplex Master Mix Kit (Thermo Fisher) was used to amplify nucleic acid according to the manufacturer’s instructions, using custom primer pools in 20 μl reactions. Primer pools of 150 nM, 300 nM, and 500 nM and anneal temperatures of 58 °C and 60 °C and were all tested and compared to determine optimal conditions. The following thermal cycling conditions were used: (1) UNG passive reference incubation at 25 °C for 2 min; (2) reverse-transcription incubation at 50 °C for 15 min; (3) enzyme activation at 95 °C for 2 min (4) 40 cycles of 95 °C for 3 s and 60 °C for 30 s. Amplified samples were directly added into the detection reaction or stored at −20 °C until use.

#### 4c. GoTaq® Probe RT-qPCR Kit (Promega)

The GoTaq® Probe RT-qPCR Kit (Promega) was used to amplify nucleic acid via RT-PCR according to the manufacturer’s instructions, using custom primer pools in 20 μl reactions. Primer pools of 200 nM, 300 nM, and 500 nM were tested and compared to determine optimal conditions. Each target in the panel was amplified with its corresponding primer pool. The following thermal cycling conditions were used: (1) reverse-transcription at 45 °C for 15 min and 95 °C for 2 min; (2) 40 cycles of 95 °C for 15 s, 60 °C for 1 min. Amplified samples were directly added into the detection reaction or stored at −20 °C until use.

#### 4d. UltraPlex™ 1-Step ToughMix® (4X) (Quantbio)

The UltraPlex™ 1-Step ToughMix® (4X) (Quantbio) was used to amplify nucleic acid via RT-PCR according to the manufacturer’s instructions, using custom primer pools in 20 μl reactions. Primer pools of 200 nM, 300 nM, and 500 nM were tested and compared to determine optimal conditions. Each target in the panel was amplified with its corresponding primer pool. The following thermal cycling conditions were used: (1) reverse-transcription at 50 °C for 10 min and 95 °C for 3 min; (2) 45 cycles of 95 °C for 10 s, 60 °C for 1 min. Amplified samples were directly added into the detection reaction or stored at −20 °C until use.

### 5. RVP Testing at Broad research laboratory (Fig. 2)

#### 5a. Design of 9-virus respiratory panel and RNase P

We designed this panel according to the methods described above under ‘Respiratory Panel - Design’ for these 9 viruses: SARS-CoV-2, HCoV-HKU1, HCoV-OC43, HCoV-NL63, FLUAV, FLUAV-g4, FLUBV, HPIV-3, HRSV, and HMPV, with the addition of an RNase P primer pair and crRNA. RNase P primers and crRNAs were designed within the same region of the gene as the CDC RT-qPCR assay (Supp. Table 2).

#### 5b. Patient Specimen Validation

All patient specimens evaluated on RVP were additionally evaluated concurrently with the CDC 2019-nCoV Real-Time RT-PCR Diagnostic Panel for N1 and RNase P. A subset of specimens were selected for further study using Next Generation Sequencing.

Of the 533 patient specimens evaluated only two specimens had no detectable levels of RNase P above threshold, one of which was positive for SARS-CoV-2 while the other was virus-negative. The RNase P negative, but virus-positive, specimen likely has a high concentration of viral RNA, which sequesters amplification materials during the reaction, limiting RNase P amplification. The double negative specimen suggests possible extraction failure or sample integrity issues and was thus excluded from further analysis.

##### 5bi. RVP Detection

Specimen preparation was performed according to the method outlined in ’General mCARMEN Procedures - Sample Extraction’ with 200 µL of input material. Amplification was performed according to methods outlined in ’General mCARMEN Procedures - Amplification’ Detection reactions were prepared as described in ‘General mCARMEN Procedures - Detection’ with the following modifications: 42.5 nM LwaCas13 and 212.5 nM crRNA in each assay mix reaction, and 500 nM quenched synthetic fluorescent RNA reporter (FAM/rUrUrUrUrUrUrU/3IABkFQ/) in each sample mix reaction. Results were analyzed following methods outlined under ‘General mCARMEN Procedures - Data Analysis’.

##### 5bii. CDC 2019-nCoV Real-Time RT-PCR Diagnostic Panel - Research Use Only

The CDC 2019-nCoV Real-Time RT-PCR Diagnostic Panel was performed using TaqPath 1-Step RT-qPCR Master Mix (Thermo Fisher) with 1 μL template RNA of either SARS-CoV-2 or RNase P in 10 μL reactions, run in triplicate. Primers from the <u>2019-nCoV RUO Kit</u> (IDT) were used. For SARS-CoV-2 a primer pool at 800 μM and probe at 200 μM was used. For RNaseP, a primer pool at 500 μM and a probe at 125 μM was used. The following thermal cycling conditions were used: (1) enzyme activation at 25 °C for 2 min; (2) reverse-transcription at 50 °C for 15 min; (3) PCR activation at 95 °C for 2 min; (4) 45 cycles of 95 °C for 3 s and 55 °C for 30 s. Standard curves were made with spike-in of RNA template (SARS-CoV-2 and RNaseP) to make a ten-fold serial dilution from 10^0^ to 10^6^ copies/μL. This was run on the QuantStudio™ 6 Flex System.

##### 5eiii. Next Generation Sequencing

Metagenomic sequencing libraries were generated as previously described ^6,50^. Briefly, extracted RNA was DNase treated to remove residual DNA then human rRNA was depleted. cDNA was synthesized using random hexamer primers. Sequencing libraries were prepared with Illumina Nextera XT DNA library kit and sequence with 100-nucleotide or 150-nucleotide paired-end reads. Data analysis was conducted on the Terra platform (app.terra.bio); all workflows are publicly available on the Dockstore Tool Registry Service. Samples were demultiplexed using demux_plus to filter out known sequencing contaminants. SARS-CoV-2 genomes were assembled using assemble_refbased, all other viral genomes were assemble_denovo and additionally visualized using classify_kraken, geneious, and R. A virus was determined to be present if more than 10 reads mapped to a particular viral genome. Full genomes were deposited into GenBank (Accession # here).

### 6. Clinical Evaluation of RVP in CLIA-certified laboratory at MGH (Fig. 3)

#### 6a. RVP preparation

##### 6ai. Design

We designed this panel according to the methods described above under ‘RVP Testing at Broad - Design’.

##### 6aii. Controls

*Automated Extraction, KingFisher Control: Extraction Negative Control (EC)* is an RNA extraction control and is prepared by adding 200 μL pooled human sample (negative for all viruses on the panel) to a well with 280 μL of binding bead mix (preparation described in ’Automated Extraction, KingFisher’). The EC should yield a positive result for the RNaseP crRNA and primer pair and a negative result for all other targets.

*Nucleic acid amplification controls: No Template Control (NTC)* is a negative control for nucleic acid amplification and is prepared by adding 10 μL nuclease-free water (instead of RNA) into 40 μL of OneStep RT-PCR Kit (Qiagen) mastermix. This should yield a negative result for all targets on the panel. *Combined Positive Control (CPC)* is a positive control for nucleic acid amplification and is prepared by pooling *in vitro* transcribed synthetic RNA of all the targets on the panel to 10^3^ copy/µl. 11 µL aliquots of this mix are stored at -80°C until use, when 10 μL are added to 40 μL of OneStep RT-PCR Kit (Qiagen) mastermix. This should yield a positive result for all targets on the panel.

*Cas13-Detection Controls: Negative Detection Control (NDC)* is a negative control for the Fluidigm-detection step and is prepared by adding nuclease-free water (instead of amplified RNA) to the sample mix without MgCl_2_. This should yield a negative result for all targets on the panel. *No crRNA control (no-crRNA)* is a negative control for the Fluidigm-detection step and is prepared by adding nuclease-free water (instead of 1 µM crRNA) to the assay mix. This should yield a negative result for all targets on the panel.

##### 6aiii. Batch preparation of sample and assay mixtures

Sample and assay mixtures can be prepared in advance for multiple 96-sample batches following similar methods, with the following changes: the batch sample mix contained all reagents described above, excluding 9 mM MgCl_2_, and the batch assay mix contained all reagents described above, excluding 2x Assay Loading Reagent. Both mixtures were calculated with 10% overage. Both mixtures were stored at −80 °C until use. 9 mM MgCl_2_ was added to the sample mix and 2x Assay Loading Reagent was added to each assay mix before use.

##### 6aiv. SYBR RT-qPCR of viral seed stock and genomic RNA from ATCC and BEI Resources

Quantification of all viral seed stock and genomic RNA received from ATCC and BEI resources was performed using the Power SYBR Green RNA-to-Ct 1-Step Kit (ThermoFisher). Reactions were run in triplicate with 1 μL RNA input in 10 μL reactions. A primer mix at 500nM was used, and all primer sequences used are listed in Supp. Table 2. The following thermal cycling conditions were used: (1) reverse transcription at 48 °C for 30 min; (2) enzyme activation at 95 °C for 10 min; (3) 40 cycles of 95 °C for 15 s and 60 °C for 1 min; (4) melt curve of 95 °C for 15 s, 60 °C for 15 s, and 95 °C for 15 s. Standard curves were made with spike-in of RNA template to make a ten-fold serial dilution from 10^0^ to 10^6^ copies/μL. This was run on the QuantStudio™ 6 Flex System.

#### 6b. Limit of Detection (LOD)

Samples were prepared for the LOD experiments using either quantified viral isolates, genomic RNA or IVT partial gene fragments. For the SARS-CoV-2, HCoV-OC43, HRSV, and HPIV-3 assays, quantified viral isolates of known titer (RNA copies/µL) spiked into pooled negative human sample (negative for all viruses on the panel) in Universal Transport Media (UTM), to mimic clinical specimen. The pooled human negative samples were incubated in the binding bead mix solution according to the methods described in ‘General mCARMEN Procedures - Extraction - Automated.’ Since no quantified virus isolates for HCoV-NL63, HCoV-HKU-1, FLUAV, FLUAV-g4, FLUBV, and HMPV were available for use at the time the study was conducted, assays designed for RNA detection of these viruses were tested with either genomic RNA from ATCC (FLUAV: cat# NR-43756; FLUBV: cat# VR-1804) or IVT RNA of known titer were spiked into pooled negative human samples in UTM.

RNA was extracted from 200 µL of input material using the MagMAX™ DNA Multi-Sample Ultra 2.0 Kit on a KingFisher™ Flex Magnetic Particle Processor with 96 Deep-Well Head (Thermo Fisher Scientific). This was run according to the protocol listed in <u>TaqPath™ COVID-19 Combo Kit Protocol</u> under “KingFisher, “Extract RNA - Automated method (200 μL input volume)” with the following differences: To prepare the binding bead mix, the following was added: 265 μL binding solution, 10 μL total nucleic acid magnetic beads, 5 μL Proteinase K with 10% overage for multiple samples. 280 μL of the binding bead mix was added to each sample well. The 200 μL of input material includes negative human sample and RNA: 160 μL of pooled human samples (negative for all viruses on the panel) was added to each sample well and incubated for 20 minutes before 40 μL of RNA was spiked in. Samples were eluted in 50 μl of elution solution and either directly added to the amplification reaction or stored at −80 °C until use.

A preliminary LOD for each assay was determined by testing triplicates of RNA purified using the extraction method described in ’RVP panel’. The approximate LOD was identified by extraction, amplification, and detection of 10-fold serial dilutions of IVT RNA of known titer (copies/µL) for 5 replicates. These concentrations ranged from 1e4-1e-3 copies/μL. The lower bound of the LOD range was determined as the lowest concentration where 5/5 replicates were positive, and the upper bound was determined as the concentration 10-fold above the lower bound. A confirmation of the LOD for each assay was determined by testing 20 replicates of RNA, purified using the extraction method described in ’RVP panel’. The approximate LOD was identified by extraction, amplification, and detection of 2-fold serial dilutions of the input sample, quantified viral isolates, genomic RNA or IVT RNA. These concentrations ranged from 20-0.5 copies/μL, depending on the virus. The LOD was determined as the lowest concentration where ≥95% (19/20) of the replicates were positive.

#### 6c. Specificity

##### Design of targets for specificity

###### In silico analysis - Inclusivity

Inclusivity was tested by performing an *in silico* analysis using all publicly available sequences of all targets on the RVP panel. Complete genomes for all viruses were downloaded from NCBI on April 2, 2021 and aligned using MAFFT v7. For viral species with less than 1000 sequences, the FFT-NS-ix1000 algorithm was used to create the MAFFT alignment. For viral species with >1000 sequences, the FFT-NS-1 algorithm was used to create the MAFFT alignment. The primer and crRNA sequences were then mapped to the aligned viral sequences using a consensus alignment to determine the percent identity (homology) and the number of mismatches. The average homology and mismatches were taken across the total number of sequences evaluated. Please note that mismatches below for crRNA sequences do not take wobble base pairing (G-U pairing) into account. Results are summarized in Supp. Table 5.

Additionally the SARS-CoV-2 crRNA and primer sequences were tested by NCBI BLAST+ against the nr/nt databases (updated 03/31/2021, N=68965867 sequences analyzed) and the Betacoronavirus database (updated 04/01/2021, N=140760). The search parameters were adjusted to blastn-short for short input sequences. The match and mismatch scores are 1 and -3, respectively. The penalty to create and extend a gap in an alignment is 5 and 2, respectively. Blast results confirmed only perfect matches to SARS-CoV-2.

###### In silico analysis - Specificity

Complete genomes for all viruses were downloaded from NCBI on April 2, 2021 and aligned using MAFFT. For viral species with less than 1000 sequences, FFT-NS-ix1000 was used. For viral species with >1000 sequences, FFT-NS-1 was used for the MAFFT alignment. The primer and crRNA sequences were then mapped to the aligned viral sequences using a consensus alignment to determine percent identity (homology). The average homology was taken across the panel sequences and the total number of sequences evaluated. Bolded text represents on-target primers/crRNA to the intended viral sequences. Not all sequence combinations were evaluated since whole genome homology between many viruses is significantly less than 80%. All primer and crRNA sequences do not have >80% homology to other, unintended viral or bacterial sequences, making the panel highly specific to particular viruses of interest. More specifically, no in silico cross-reactivity >80% homology between any primers and crRNA sequences on RVP is observed for the following common respiratory flora and other viral pathogens: SARS-CoV-1, HCoV-MERS, Adenovirus, Enterovirus, Rhinovirus, *Chlamydia pneumoniae, Haemophilus influenzae, Legionella pneumophila, Mycobacterium tuberculosis, Streptococcus pneumoniae, Streptococcus pyogenes, Bordetella pertussis, Mycoplasma pneumoniae, Pneumocystis jirovecii, Candida albicans, Pseudomonas aeruginosa, Staphylococcus epidermis, Streptococcus salivarius*. In silico analysis results are summarized in Supp. Table 6.

###### In vitro analysis

Samples were prepared for specificity experiments according to methods described above in ‘General mCARMEN Procedures - Preparation of IVT material’, and samples were serially diluted down to a concentration of 10^6^ and 10^5^ copies/µL. For all samples prepared for specificity experiments, RNA was extracted from 200 µL of input material using the MagMAX™ DNA Multi-Sample Ultra 2.0 Kit on a KingFisher™ Flex Magnetic Particle Processor. This was run according to the extraction, amplification, and detection methods described above under ‘RVP Testing at Broad.’

#### 6d. Patient Specimen Validation

##### 6di. Specimen preparation prior to extraction

All patient specimens from the MGH Clinical Microbiology Lab were initially reported to be positive for HCoV-HKU1, HCoV-NL63, and HMPV via BioFire FilmArray Respiratory Panel (RP2) or positive for SARS-CoV-2, FLUAV (H3), FLUBV, and HRSV via <u>Xpert® Xpress SARS-CoV-2/Flu/RSV (Cepheid)</u>. 200 µL of positive for SARS-CoV-2 were aliquoted as follows: 220 μL for testing using the RVP panel, 220 μL for testing using the TaqPath™ COVID-19 Combo Kit, and all remaining specimens were stored at −80 °C. All negative specimens were aliquoted: 220 μL for RVP panel testing, 220 μL for TaqPath™ COVID-19 Combo Kit testing, 400 μL for BioFire FilmArray Respiratory Panel (RP2) testing, and all remaining specimen was stored at −80 °C. All other specimens were aliquoted: 220 μL for RVP panel testing, 400 μL for BioFire FilmArray Respiratory Panel (RP2) testing, and all remaining specimen was stored at −80 °C.

##### 6dii. Preparation of contrived samples prior to extraction

Contrived patient samples of viruses HCoV-HKU1, HCoV-OC43, HCoV-NL63, FLUAV-g4, HPIV-3, and HMPV were prepared by diluting either viral seed stock (HCoV-OC43 and HPIV-3) or template RNA (HCoV-HKU1 and HCoV-NL63). See Supp. Table 7 for viral seed stock vendor details. The viral seed stock or template RNA is added to non-pooled human specimens (negative for all targets, except RNase P, on the RVP panel) at a concentration 2 times the LOD; the concentrations for these samples ranged from 10^2^ to 10^6^ copies/µL.

##### 6diii. RVP

All materials were extracted, amplified, detected, and analyzed using methods described under ‘General mCARMEN Procedures ’ and ‘RVP Testing at Broad - Patient Specimen Validation.’

##### 6div. TaqPath™ COVID-19 Combo Kit

A subset of patient specimens, all SARS-CoV-2 and negative patient specimens, from the MGH Clinical Microbiology Laboratory were verified using the TaqPath™ COVID-19 Combo Kit (ThermoFisher). These samples were initially reported to be positive for SARS-CoV-2 via Xpert® Xpress SARS-CoV-2/Flu/RSV (Cepheid) or reported to be negative for all targets (excluding RNaseP) on the RVP panel via BioFire FilmArray Respiratory Panel (RP2).The TaqPath™ COVID-19 Combo Kit was performed according to the manufacturer’s instructions. The assay was performed using the Applied Biosystems 7500.

##### 6dv. BioFire FilmArray Respiratory Panel (RP2)

A subset of patient specimens from the MGH Clinical Microbiology Laboratory, all HCoV-HKU1, HCoV-NL63, FLUAV (H3), FLUBV, HRSV, HMPV and negative patient specimens, were verified using the BioFire FilmArray Respiratory Panel (RP2) (Biofire Diagnostics). These specimens were either initially reported to be positive for HCoV-HKU1, HCoV-NL63, and HMPV via BioFire FilmArray Respiratory Panel (RP2) or positive for FLUAV (H3), FLUBV, and HRSV via <u>Xpert® Xpress Flu/RSV (Cepheid)</u>. For each run, one patient specimens in UTM at 300 µL was verified using the BioFire FilmArray Respiratory Panel (RP2) according to the <u>manufacturer’s instructions</u>. Any remaining specimen was stored at -80°C.

Controls for this assay were received with the kit and ready for use. Control 1 is expected to be positive for adenovirus, HMPV, Human Rhino/Enterovirus, FLUAV (H1-2009), FLUAV (H3), HPIV-1, HPIV-2. Control 2 is expected to be positive for HCoV-229E, HCoV-HKU1, HCoV-NL63, HCoV-OC43, FLUAV (H1), FLUBV, HPIV-2, HPIV-3, HRSV.

The results were automatically displayed on the FilmArray software with each target in a run reported as “detected” or “not detected.” If either control fails, the software marks this run as “invalid.” When sufficient human sample volume was available, samples with invalid results were re-run.

#### 6e. Analysis Software

The analysis software comprises python scripts executing the data analysis described in Methods 2h and taking into account the controls described in Methods section 6aii. They are packaged into an executable with graphical user-interface using the Python module Gooey 1.0.7.

In short, the reference-normalized background-subtracted fluorescence is calculated for guide-target pairs for the measurement after 60 min. Then, the dynamic range and the separation band are assessed.

Separation band = (mean of positive controls - 3 standard deviations of positive controls) - (mean of negative controls - 3 standard deviations of negative controls) Dynamic range = | mean of positive controls - mean of negative controls|

If the ratio of separation band to dynamic range is equal or less than 0.2, the whole assy is invalid. Next, for the positive and negative controls, outliers based on three standard deviations are identified. If a positive control has a too low value or a negative control a too high value, the respective assay is invalid. For the remaining samples, hit calling is performed based on comparing the signal to the water control. If the signal is 1.8x higher than the water control, the guide-target pair is called a hit. Based on this hit calling, the extraction control, the negative and positive detection controls and internal controls are verified. If their result does not correspond to their expected hit status, either the respective assay or specimens is turned invalid. All patients specimens to be valid need to be either positive for RNase P or at least one other assay. Finally, the software annotates the results as csv files and visualizes them as an annotated heatmap.

### 7. Variant Testing (Fig. 4)

#### 7a. Design

The crRNAs for SNP discrimination were designed using a generative sequence design algorithm (manuscript in prep.). This approach uses ADAPT’s predictive model to predict the activity of candidate crRNA sequences against on-target and off-target sequences^35^. These predictions of candidate crRNA activity steer the generative algorithm’s optimization process, in which it seeks to design crRNA probes that have maximal predicted on-target activity and minimal predicted off-target activity. Using this design algorithm, we selected the 26 mutations to detect and discriminate between the variants (Supp. Table 2).

#### 7b. Amplification

We followed the methods under ‘General mCARMEN Procedures - Amplification - SSIV,’ sequences of primers used can be found in Supp. Table 2.

#### 7c. Detection

We followed the methods under ‘General mCARMEN Procedures - Detection’ with the following modifications: 42.5 nM LwaCas13 and 2-212.5 nM crRNA in each assay mix reaction, and 500 nM RNaseAlert v2 in each sample mix reaction. Exact crRNA concentrations can be found in Supp. Table 2.

#### 7d. Data analysis

##### Threshold calculation

To determine if an ancestral or derived sequence is present, the signals between respective ancestral and derived crRNA pairs must be evaluated and compared (Supp. Table 2). First, background subtracted reporter fluorescence is normalized to the background subtracted passive reference dye (ROX) fluorescence for each assay in the IFC. Next, the ancestral:derived and derived:ancestral ratios are calculated for each five minute-interval time point across 180 minutes. For each crRNA pair, the ratio reaching a crRNA pair-specific threshold at the earliest time point is selected. If the ancestral:derived ratio is selected, then the sequence present is determined to be ancestral. If the derived:ancestral ratio is selected, then the sequence present is determined to contain the mutation targeted by the derived crRNA within the SARS-CoV-2 spike gene. crRNA pair-specific thresholds were determined based on ancestral and variant control samples, also referred to as the seedstock samples, tested in parallel with the unknown samples. For a given crRNA pair, the threshold was set to the lowest value with the maximum combined sensitivity and specificity when applied to the seedstock samples. For crRNA’s detecting a SNP at the same position, the second lowest threshold with the maximum combined sensitivity and specificity was chosen if possible without compromising the maximum combined sensitivity and specificity. For crRNA pairs targeting mutations not represented in the variant control samples, a default crRNA pair threshold of 1.5 was set. Expected mutations for each variant lineage:

“Variant Identified” hit calling parameters:

1. If no mutations are detected, a result of “Ancestral” is returned.
2. At least one unique crRNA specific to a single SARS-CoV-2 variant must be above the fluorescence ratio threshold. If there is not one unique crRNA signal above threshold, a result of “Variant Not Identified” will be returned.
3. If two or more mutations for a given variant fall below the threshold, a result of “Variant Not Identified” will be returned. All other mutations must surpass the threshold.
4. If three or more unexpected mutations for a given variant are above threshold, a result of “Variant Not Identified” will be returned. At most, two unexpected signals can occur as long as parameters 1 and 2 are met.

If all three parameters are met, the result of “Variant Identified” will be returned. If the parameters are not met, a result of “Variant Not Identified” will be returned. Samples that contain additional derived signals that fall outside of the typical variant lineage mutation list follow the below parameters. If 1-2 unexpected signals are observed slightly above threshold yet all other signals are correct for a specific variant lineage then the unexpected signal will be disregarded and the variant call will be made on the remaining signals. If more than two unexpected signals are observed above threshold and either all other signals are correct for a specific variant lineage or are not perfectly matching a result of “Variant Uncertain” will be returned.

*We would like to note that the variant identification pipeline will need to be updated as new SARS-CoV-2 mutations and variant lineages arise for proper identification.

There are a few exceptions worth mentioning: we observed crRNAs for SNPs E484Q, P681R, N501T, and L452Q, had undesirable cross-reactive signals with a position matched or adjacent mutation, and were, thus, excluded from further evaluation.

### 8. Cas13- and Cas12-based detection with mCARMEN (Fig. 5)

#### 8a. Design for Cas12-based detection

Cas13 crRNAs from RVP were utilized. Cas12 crRNAs were manually designed in the same region of the viral genome as the Cas13 crRNAs to reduce the need for additional primer design while maintaining Cas12’s PAM requirements. Only one additional primer was designed inorder to properly amplify all targets on RVP. All crRNAs and primers are listed in Supp. Table 2.

#### 8b. Detection

We followed the methods under ‘General mCARMEN Procedures - Detection’ with the following modifications: 10-60 nM LwaCas13, 10-60 nM LbaCas12a, 125 nM Cas13a crRNA, and 125 nM Cas12 crRNA in each assay mix reaction, and 500 nM quenched synthetic fluorescent RNA reporter (FAM/rUrUrUrUrUrUrU/3IABkFQ/ and VIC/rTrTrArTrTrArTrT/3IABkFQ) in each sample mix reaction.

#### 8c. Data analysis

We generally followed the methods under ‘General mCARMEN Procedures - Analysis’ this time with also taking into account VIC signal separate from FAM signal. We used a custom python script to determine whether the FAM signal of a reaction is significantly above background by comparing it to the no-template control. If the background-subtracted and normalized fluorescence intensity is 1.8 higher than the normalized and background-subtracted no-templated control, the assay is considered positive.

## Extended Data Figures

**Extended Data Figure 1.**
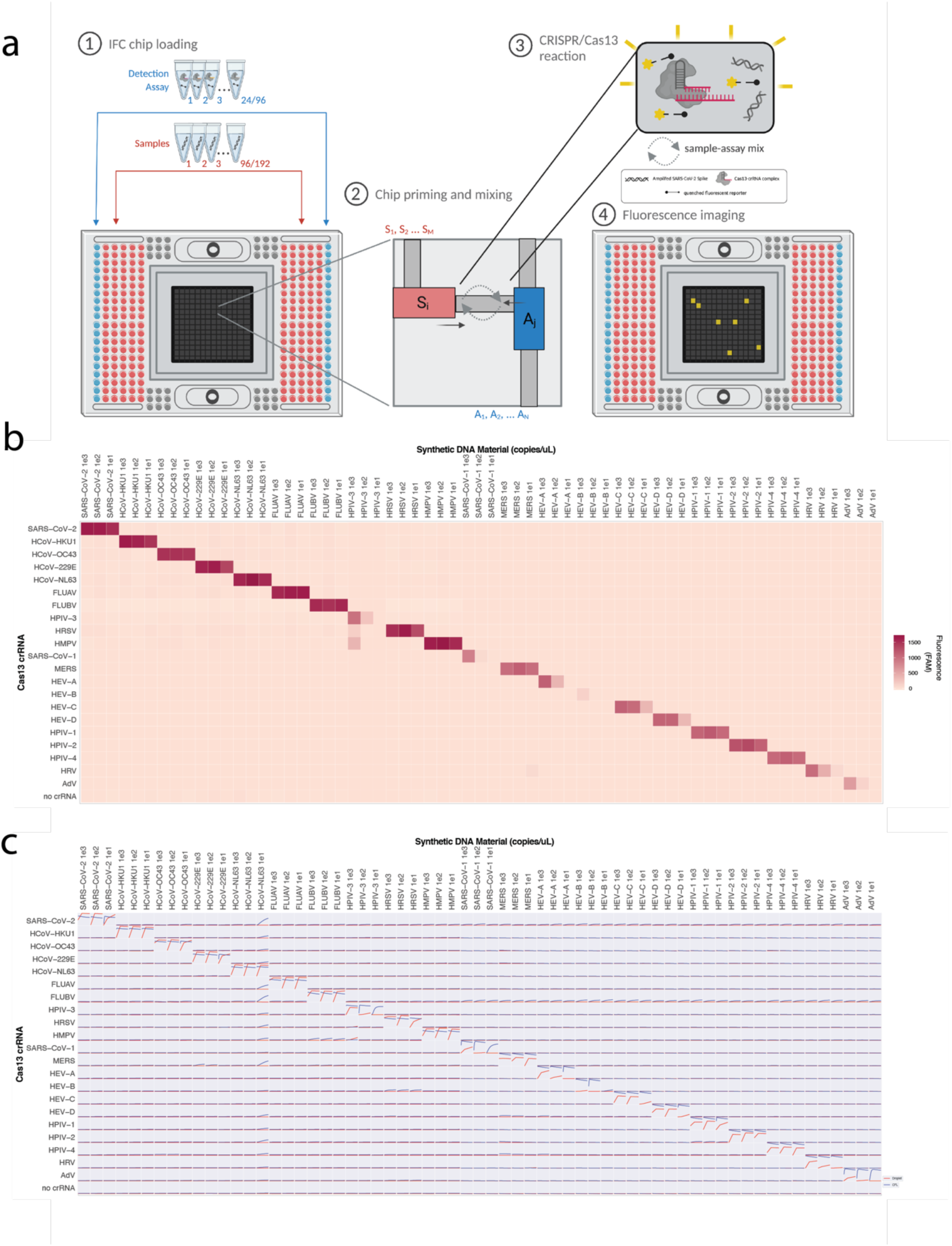
Detection of 21 respiratory viruses by mCARMEN and CARMEN v1. **a,** Detailed schematic of loading and running 192.24 IFC in mCARMEN workflow. (Step 1) Up to 196 pre-amplified samples are added in the red microwells of the Fluidigm 192.24 Dynamic Array IFC. Up to 24 detection assay reactions containing Cas13-crRNA complexes are added on the sides of the IFC shown in blue. Control line, actuation, and pressure fluids are added according to manufacturer’s guidelines. (Step 2) The IFC is loaded in a controller where the load and mix protocol is selected. In the last minute of the protocol, the individual channels open to allow sample and detection assay mixing to occur. (Step 3) Sequence specific Cas13-crRNA complexes recognize and bind to RNA which unleashes the collateral cleavage activity of Cas13. Cas13 cleaves the quenched fluorescent reporter in solution. (Step 4) The IFC is loaded into the Fluidigm Biomark after the controller protocol finishes. The Biomark, incubating at 37°C, takes images every 5 minutes for up to 3 hours to monitor fluorescence from the detection reactions. **b,** Synthetic DNA fragments from each of the 21 human respiratory viruses were serially diluted from 10^3^-10^1^ copies/μL and added to Q5 amplification master mix containing primer pool 1 or 2. After amplification, separate sample and detection assay reactions were prepared for fluorescence-based readout using CARMEN v1. CARMEN v1 values are shown as raw fluorescence (red), FAM signal at 3 hours. All samples were background subtracted from NTC-det negative control. **c,** Time series curves of mCARMEN (blue) from Fig. 1b and CARMEN v1 (red) from **b** at 0, 60, and 180 minutes.

**Extended Data Figure 2.**
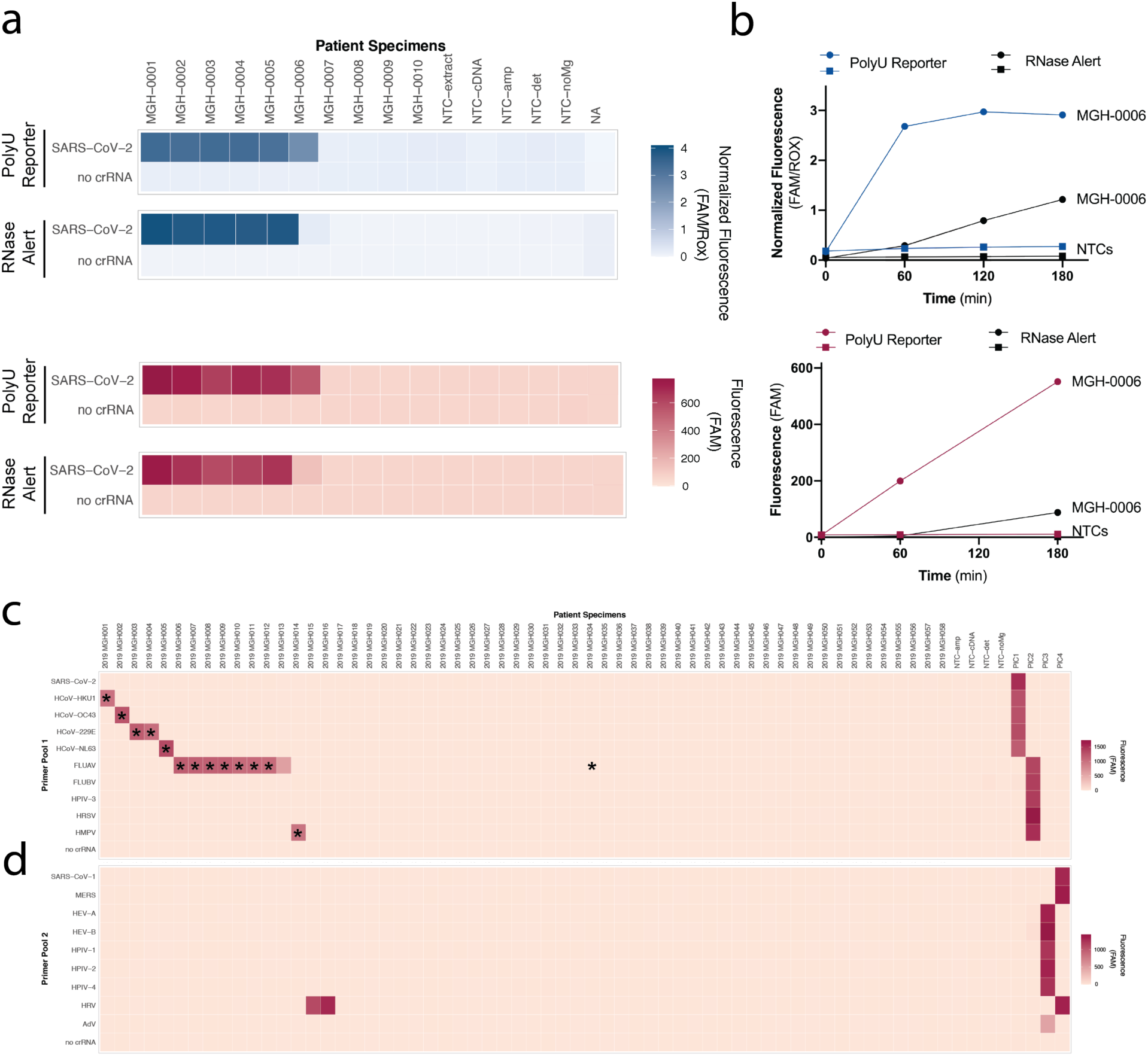
Enhanced sensitivity with polyU fluorescent reporter on patient specimens. **a,** 6 SARS-CoV-2 positive and 4 SARS-CoV-2 negative NP swabs from Figure 1e. mCARMEN values are shown as normalized fluorescence (blue) and CARMEN v1 values are shown as raw fluorescence (red) while signal using the polyU reporter is shown directly above signal using RNase Alert (Thermo). **b,** Fluorescence over time for SARS-CoV-2 positive patient specimens, MGH-006. Top, mCARMEN fluorescence values for MGH-006 and NTC with blue line representing signal from the polyU reporter and black representing signal from RNase Alert at 0, 60, 120, and 180 minutes. Bottom, CARMEN v1 fluorescence values, red line representing signal from polyU reporter and black representing signal from RNase Alert at 0, 60, and 180 minutes. **c&d,** CARMEN v1 signal shown as raw fluorescence at 3 hr post-reaction initiation. 58 patient specimens were extracted using the KingFisher Flex MagMAX mirVana kit then amplified in 2 steps with 2 separate primer pools. **c,** Amplification from primer pool 1 **d,** amplification from primer pool 2. All specimens were background subtracted using NTC-amp as a negative control. Asterisks (*) represent specimens with known infections by unbiased NGS, more than 10 reads mapped to the viral genome.

**Extended Data Figure 3.**
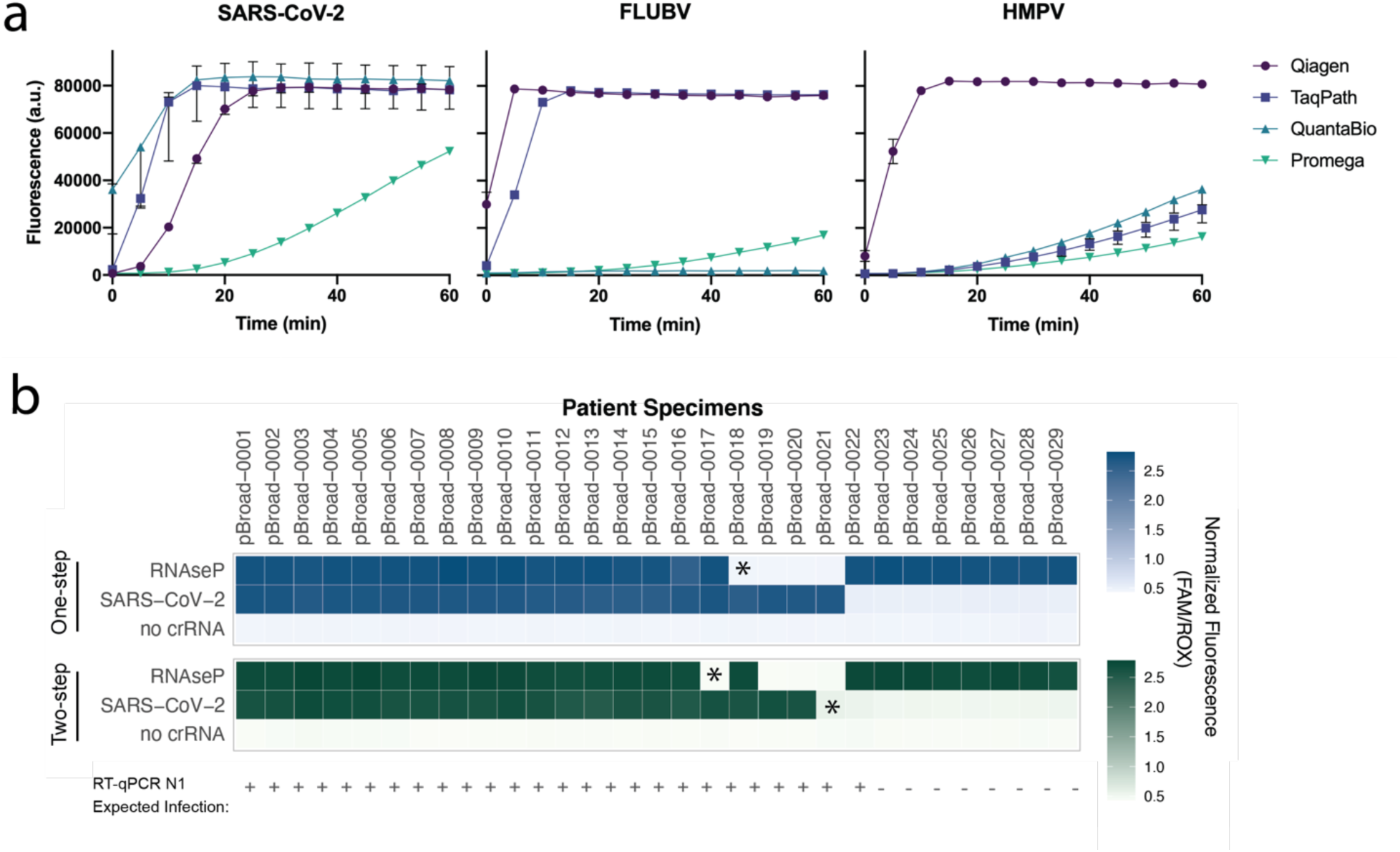
Single-step amplification troubleshooting of synthetic material and patient specimens. **a,** Fluorescence for SARS-CoV-2, FLUBV, and HMPV comparing four different single-step amplification kits Qiagen One-Step RT-PCR, TaqPath, QuantaBio, and Promega. **b,** Single-step amplification by Qiagen One-Step RT-PCR kit (top, blue) compared to two-step amplification by SSIV then Q5 (bottom, green) on 21 SARS-CoV-2 positive and 8 SARS-CoV-2 negative patient specimens. Prior RT-qPCR results shown as expected infection positive (+) or negative (-). All specimens were background subtracted using NTC-amp as a negative control. Asterisk (*) represent differences between one-step and two-step amplification results.

**Extended Data Figure 4.**
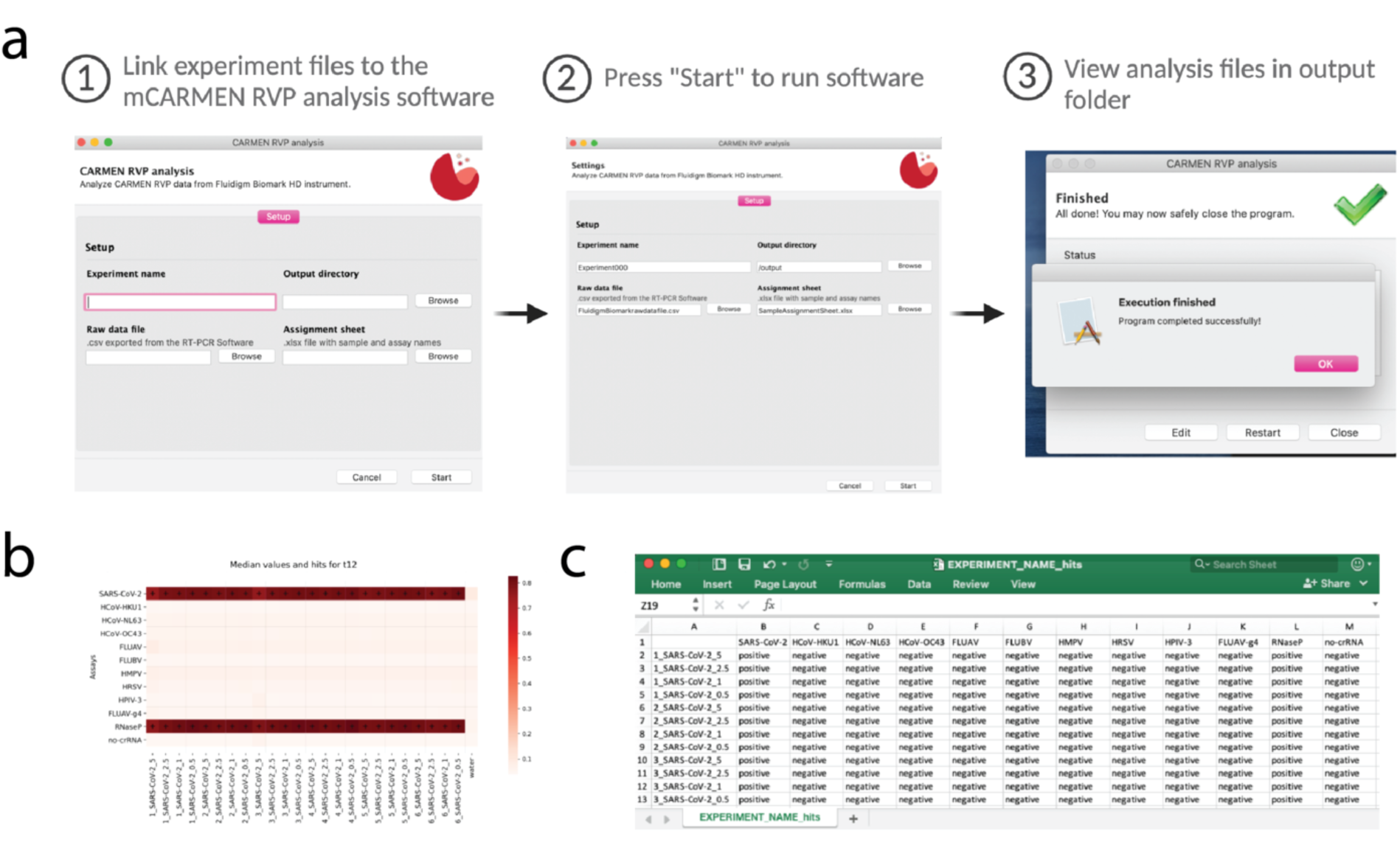
mCARMEN RVP Software for automated results and hit calling. **a,** RVP Analysis Software workflow. (Step 1) Once the software has been successfully launched, four sections must be filled in: Experiment name, Output directory, Raw data file, and Assignment sheet. The raw data file comes from the Biomark instrument after mCARMEN run is complete. The assignment sheet must be filled out prior to software analysis. (Step 2) Press start to run the RVP Software analysis. (Step 3) The analysis can take several minutes to run, but once complete the user can navigate to the specified Output directory to view the six output files. **b,** Example of a condensed heatmap from a successful mCARMEN RVP experiment ([Experiment Name_heatmap_t12.png). **c,** Snapshot of the key file that captures all the RVP results individually broken down by patient specimen ID and respiratory virus (Experiment Name_HitQuantification.csv). The file lists the Specimen IDs accompanied by a positive, negative, or invalid determination, as defined in Clinical Evaluation of “Clinical Evaluation of RVP at MGH - Controls & Software Analysis.” The CARMEN RVP Software is available at: https://github.com/broadinstitute/carmen-rvp.

**Extended Data Figure 5.**
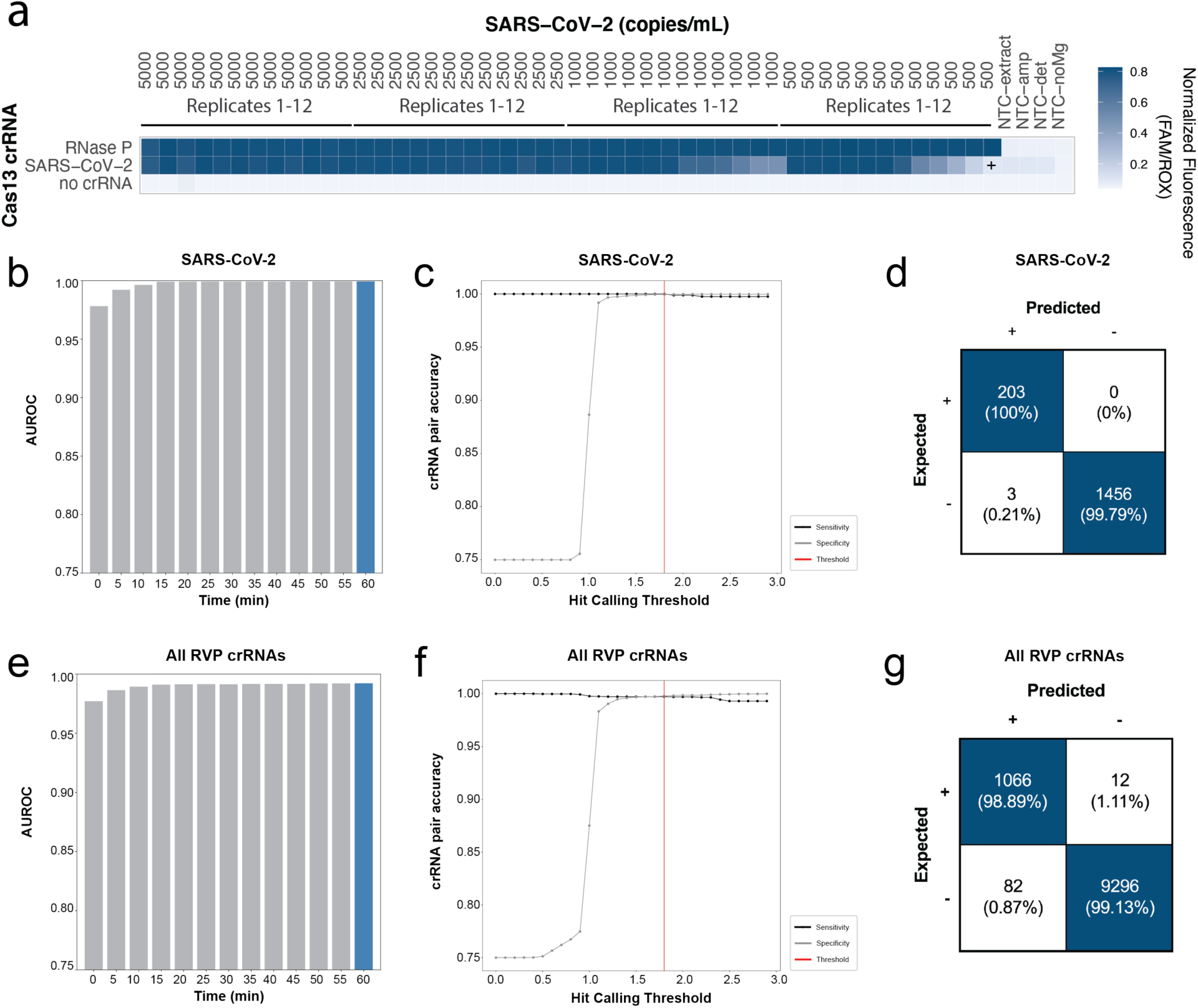
crRNA performance during RVP pre-clinical LOD evaluation. **a,** Fluorescence for 12 replicate testing of SARS-CoV-2 viral seed stock at 5,000, 2,500, 1,000, and 500 copies/mL spiked into negative patient specimens. All samples were background subtracted using NTC-amp as a negative control. Plus sign (+) represent negative sample call by mCARMEN results. **b&e,** Area under the receiver operating characteristic (AUROC) curve for crRNA detection. Blue bar represents the maximal AUROC time point at 60 minutes post-reaction initiation **c&f,** Evaluation of sensitivity and specificity of crRNA detection to establish hit calling threshold. Black line: sensitivity/true positive rate; gray line: specificity/true negative rate; red line: hit calling threshold. **d&g,** Confusion matrix of expected vs predicted detection. **b-d,** SARS-CoV-2 crRNA detection only. **e-g,** all nine respiratory viruses on RVP.

**Extended Data Figure 6.**
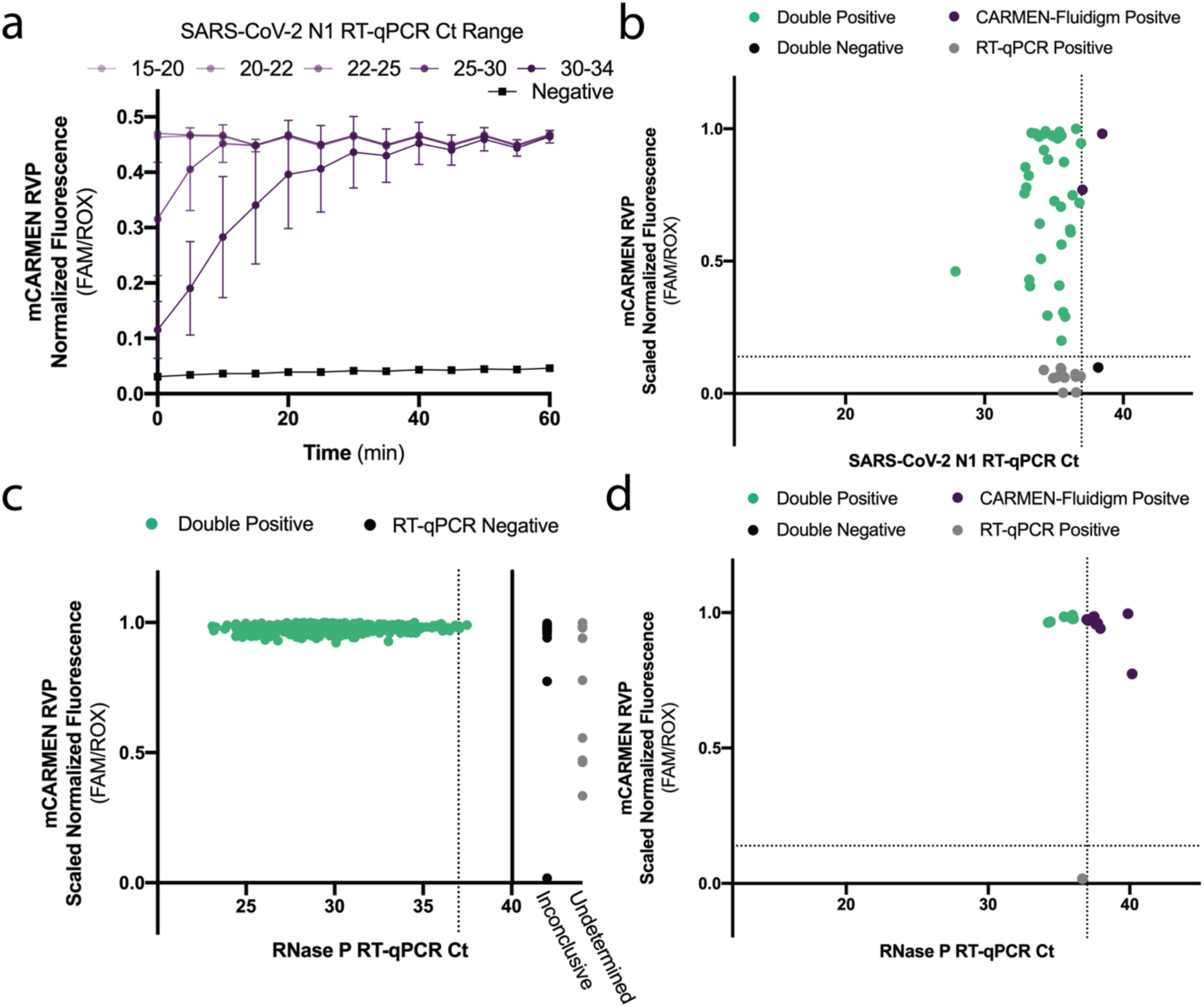
SARS-CoV-2 clinical specimen testing in an academic setting at the Broad. **a,** Average normalized fluorescence for SARS-CoV-2 across 8-14 patient specimens within Ct ranges of 15-20, 20-22, 22-25, 25-30, and 30-34. Purple lines: known SARS-CoV-2 positive specimens; black line: known SARS-CoV-2 negative specimens; error bars: one standard deviation from the mean fluorescence. **b,** SARS-CoV-2 N1 RT-qPCR inconclusive specimens from Fig. 2c. Scatter plot of mean Ct of one or two technical replicates compared to mCARMEN scaled normalized fluorescence. **c,** Scatter plot of RNase P scaled normalized fluorescence from Fig. 1b compared to RNase P Ct values obtained from concurrent RT-qPCR testing of 533 patient specimens. Green: positive RNase P signal detected by both RVP and RT-qPCR; gray: inconclusive RT-qPCR result indicating one or two of the three technical replicates were undetermined; black: undetermined RT-qPCR result indicating all three technical replicates were negative for SARS-CoV-2. **d,** RNase P RT-qPCR inconclusive specimens from **c**. Scatter plot of mean Ct of one or two technical replicates compared to mCARMEN scaled normalized fluorescence. Green: positive detection by both RVP and RT-qPCR; purple: RVP positive; gray: RT-qPCR positive; black: negative by both RVP and RT-qPCR. Dashed horizontal lines: threshold for RVP positivity. Dashed vertical line: Ct value of 37 (FDA positivity cutoff).

**Extended Data Figure 7.**
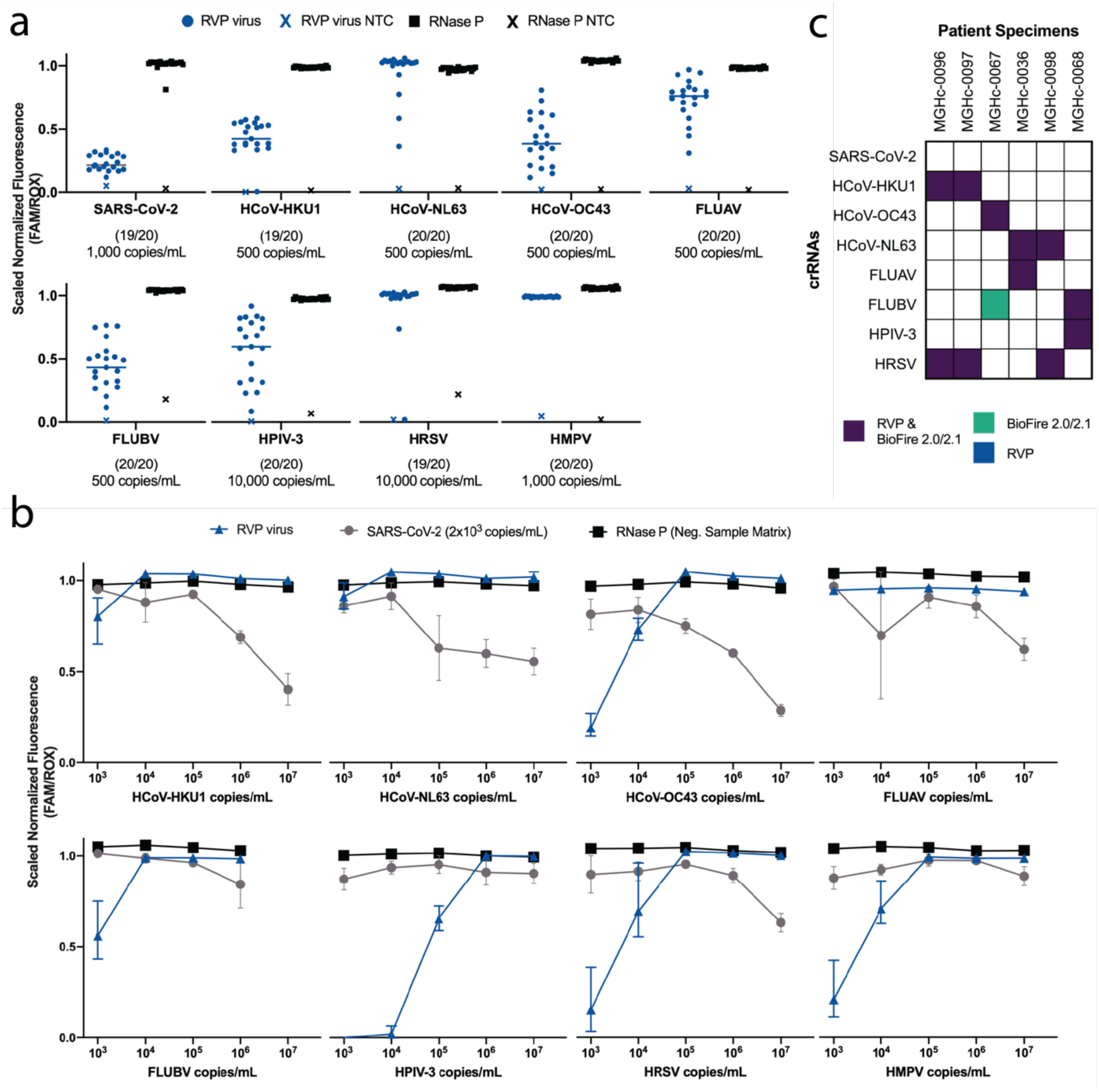
mCARMEN RVP sensitivity evaluation, LOD and co-infections, at MGH. **a,** Fluorescent values at the LOD of all 9 viruses on RVP as established by spiking viral seed stock, isolated RNA extracted from clinical specimens, or synthetic RNA into confirmed negative patient specimen for 20 replicates. NTC, no template control; Viral scaled normalized fluorescence values, circles; RNase P values, squares; NTC-noMg values, “X”. **b,** Scaled normalized fluorescence values at 1 hr are shown for SARS-CoV-2 and RNase P, as well as another RVP virus. SARS-CoV-2 viral seed stock was spiked into pooled negative specimen at 2,000 copies/mL (2x the LOD established in Fig. 3b). Additionally, viral seed stock, isolated RNA extracted from clinical specimens, or synthetic RNA was additionally spiked at 10^7^-10^3^ copies/mL. Error bars, one standard deviation from the median fluorescence. Blue: RVP virus at varying concentrations; light purple: SARS-CoV-2; black: RNase P from negative patient specimen. **c,** Concordance between RVP and Biofire RP 2.01/2.1 for 6 patient specimens with co-infections visualized as a split heatmap. Purple: RVP and BioFire 2.0/2.1 positive; blue: RVP virus-positive; green: Biofire RP2.0/2.1 virus-positive; white: virus-negative.

**Extended Data Figure 8.**
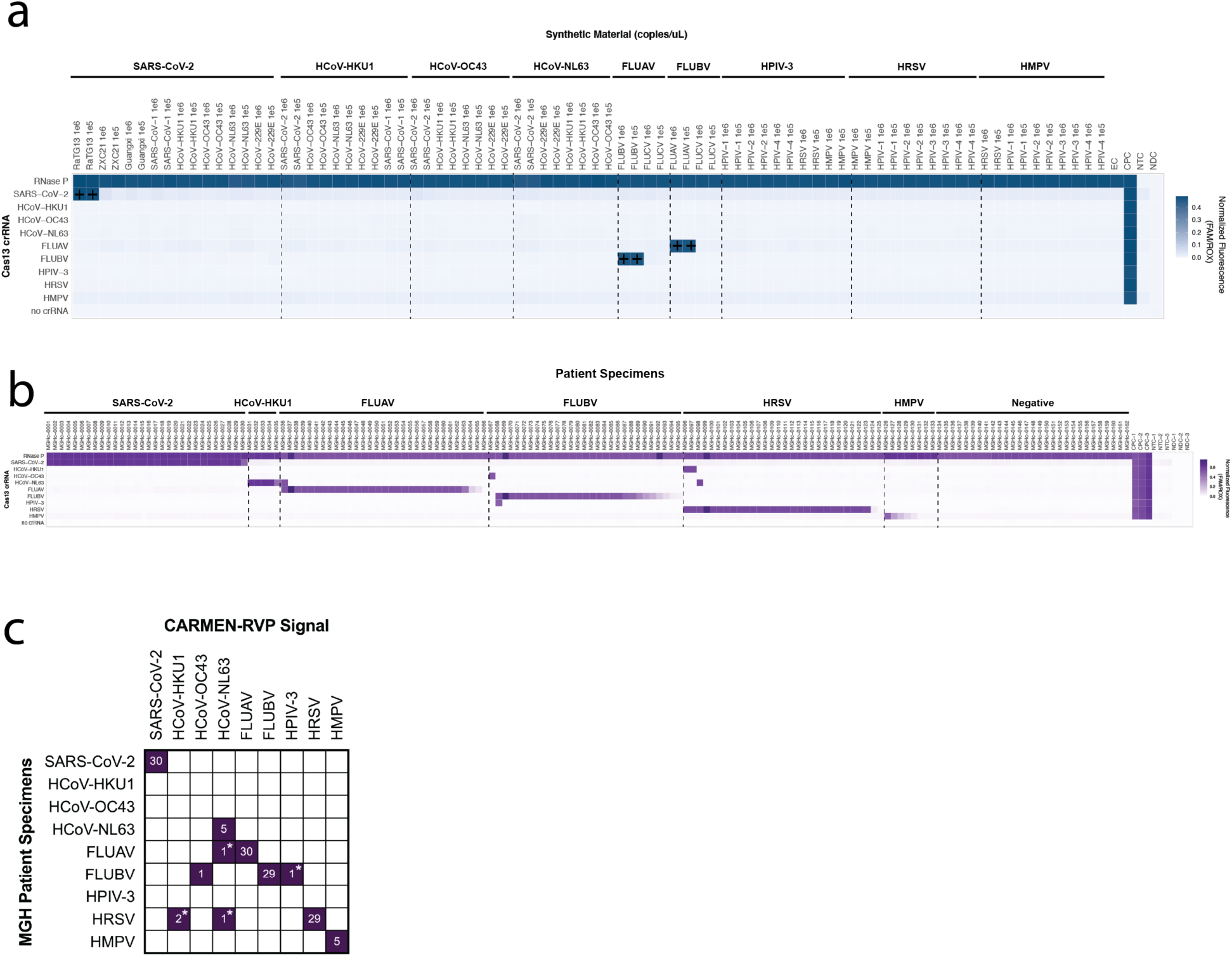
mCARMEN RVP specificity testing at MGH on *in silico* designed synthetic targets and patient specimens. **a,** Fluorescence at 1 hr post-reaction initiation on *in silico* designed synthetic targets spiked into pool negative patient specimen at 10^6^ copies/μL for specificity evaluation. Plus sign (+) represent expected cross-reactive signal. Dashed vertical line for visual aid. **b**, Fluorescence at 1 hr for 166 patient specimens with known infection status from Fig. 3d-f. Dashed vertical line for visual aid. **c**, Patient specimen summary from **b** comparing mCARMEN RVP and comparator assay results (Cepheid and BioFire 2.0/2.1). Asterisk (*) represents co-infections that were confirmed by BioFire 2.0/2.1.

**Extended Data Figure 9.**
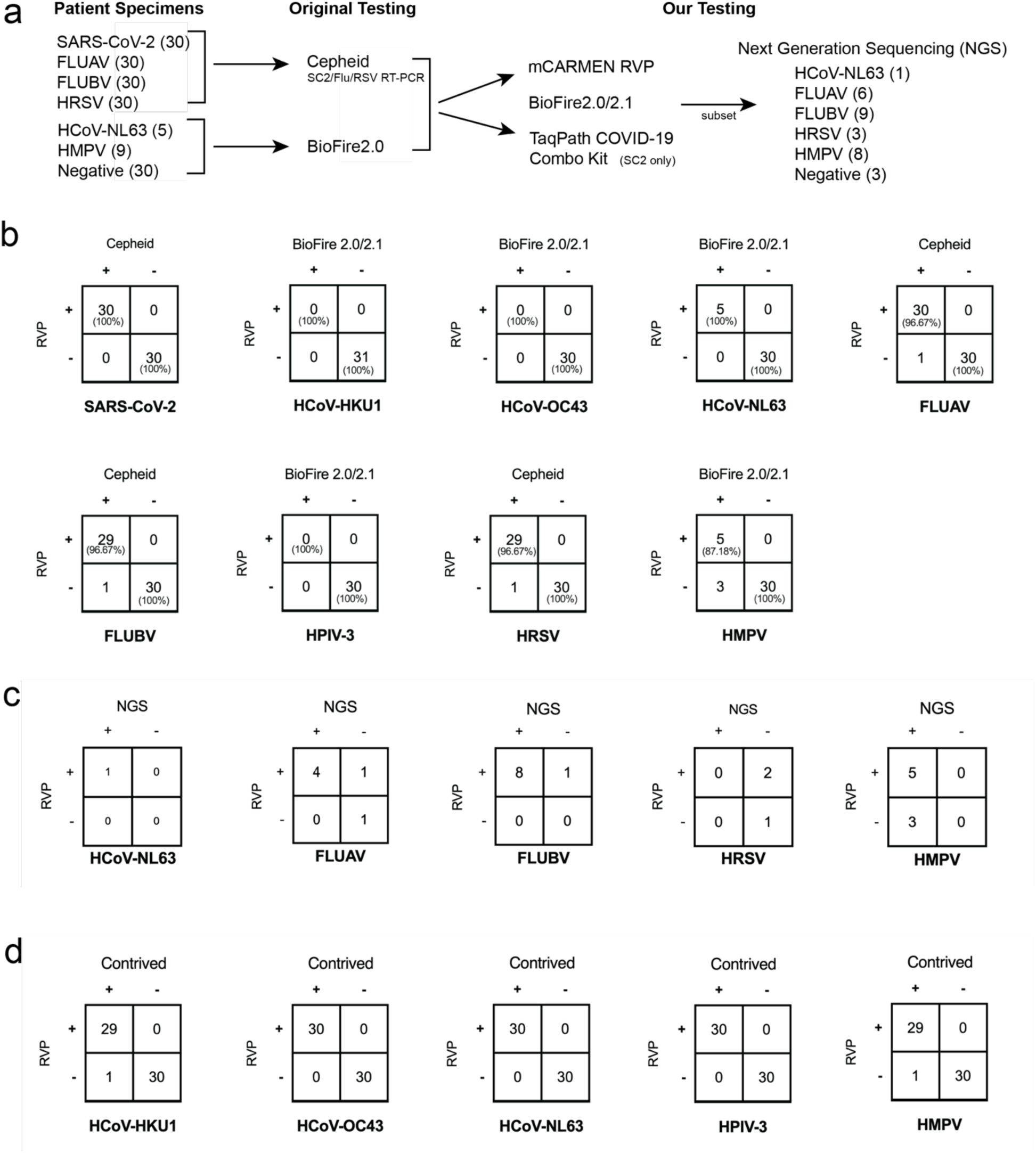
Agreement of mCARMEN patient and contrived samples tested in CLIA-certified laboratory at MGH. **a,** Workflow of patient specimens evaluation. Patient specimens were originally tested by either Cepheid and BioFire RP2.0/2.1. We tested these specimens by both mCARMEN RVP and BioFire RP2.0/2.1 then a subset of these specimens were subjected to unbiased NGS. **b,** Concordance between RVP and either Cepheid or BioFire RP2.0/2.1 for 166 patient specimens. Positive and negative percent agreements are shown in parenthesis. **c,** Concordance between RVP and NGS for 30 out of the 166 patient specimens. **d,** Concordance of contrived samples to RVP. Contrived samples were prepared by spiking viral seed stock, genomic RNA, or synthetic RNA into negative patient specimen.

**Extended Data Figure 10.**
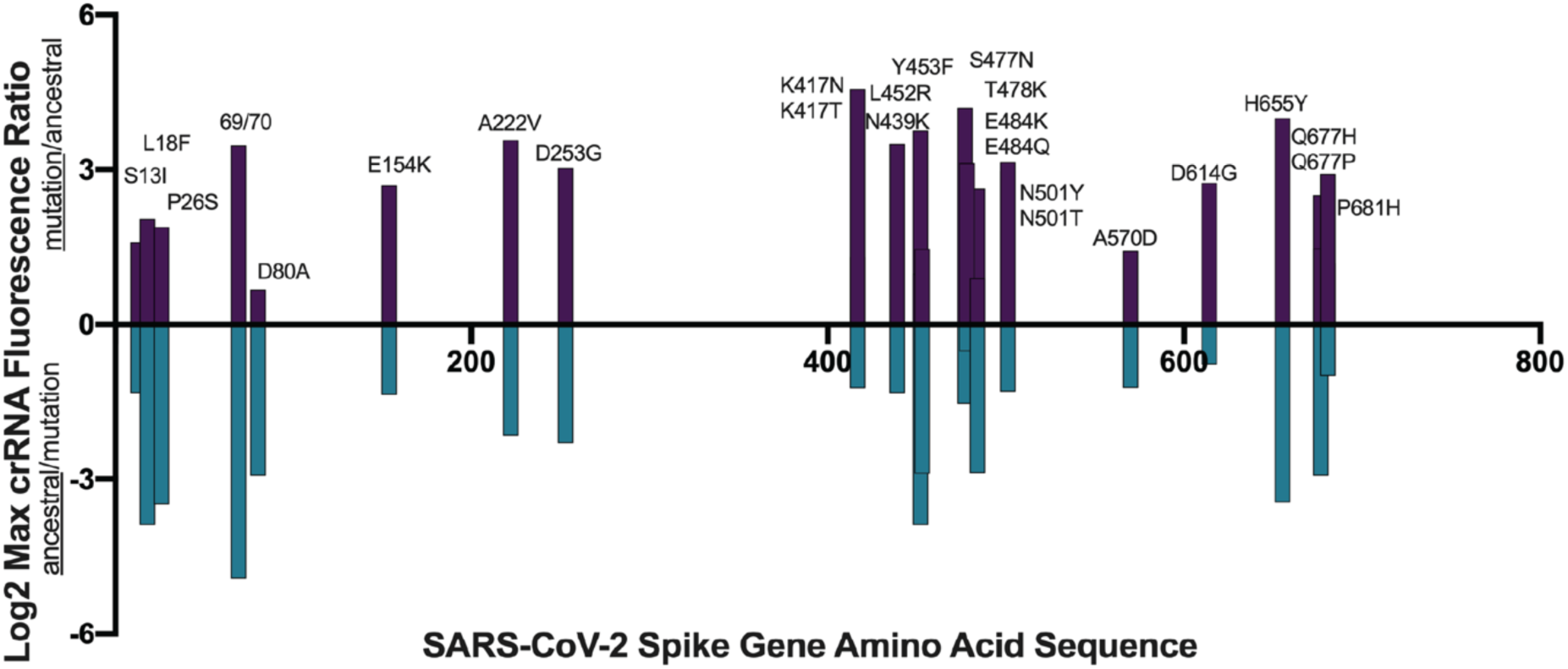
Evaluation of crRNA pairs detecting SARS-CoV-2 spike gene mutations on synthetic targets. 26 mutations across the SARS-CoV-2 spike gene were selected based on uniqueness within a variant lineage or due to phenotypic effects on viral fitness. *In vitro* transcribed synthetic RNA targets at 10^13^ copies/mL were used as input. Top, positive values, represents signal on the derived sequence (mutation/ancestral) and bottom, negative values, represents signal on the ancestral sequence (ancestral/mutation). Bars represent log_2_ maximum crRNA normalized fluorescence ratio at any 5 minute time point from 0-180 minutes after reaction initiation.

**Extended Data Figure 11.**
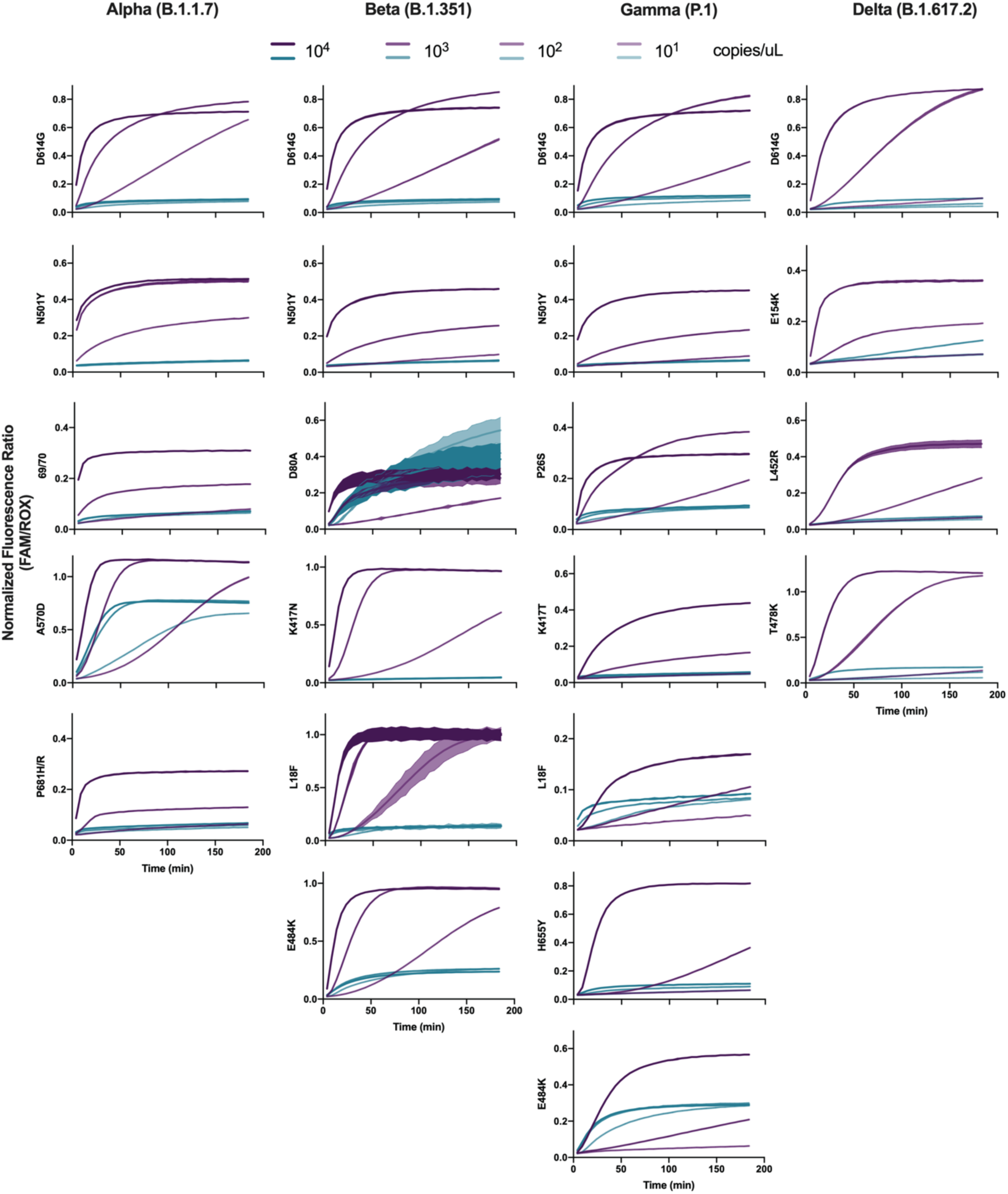
Mutation detection for Alpha, Beta, Gamma, and Delta SARS-CoV-2 variant lineages. Kinetics curves of 4-7 SNPs that make up the four different variant lineage seed stocks being evaluated. Data shown as normalized fluorescence every five minutes for 3-4 concentrations of viral seed stock. WA, Alpha, Beta, Gamma: 10^4^-10^2^ copies/μL, and Delta: 10^3^-10^1^ copies/μL. Purple: variant lineage; Blue: ancestral seed stock; colored from high-to-low concentration. Shaded regions represent 95% confidence intervals around the median fluorescence values and lines are the median.

**Extended Data Figure 12.**
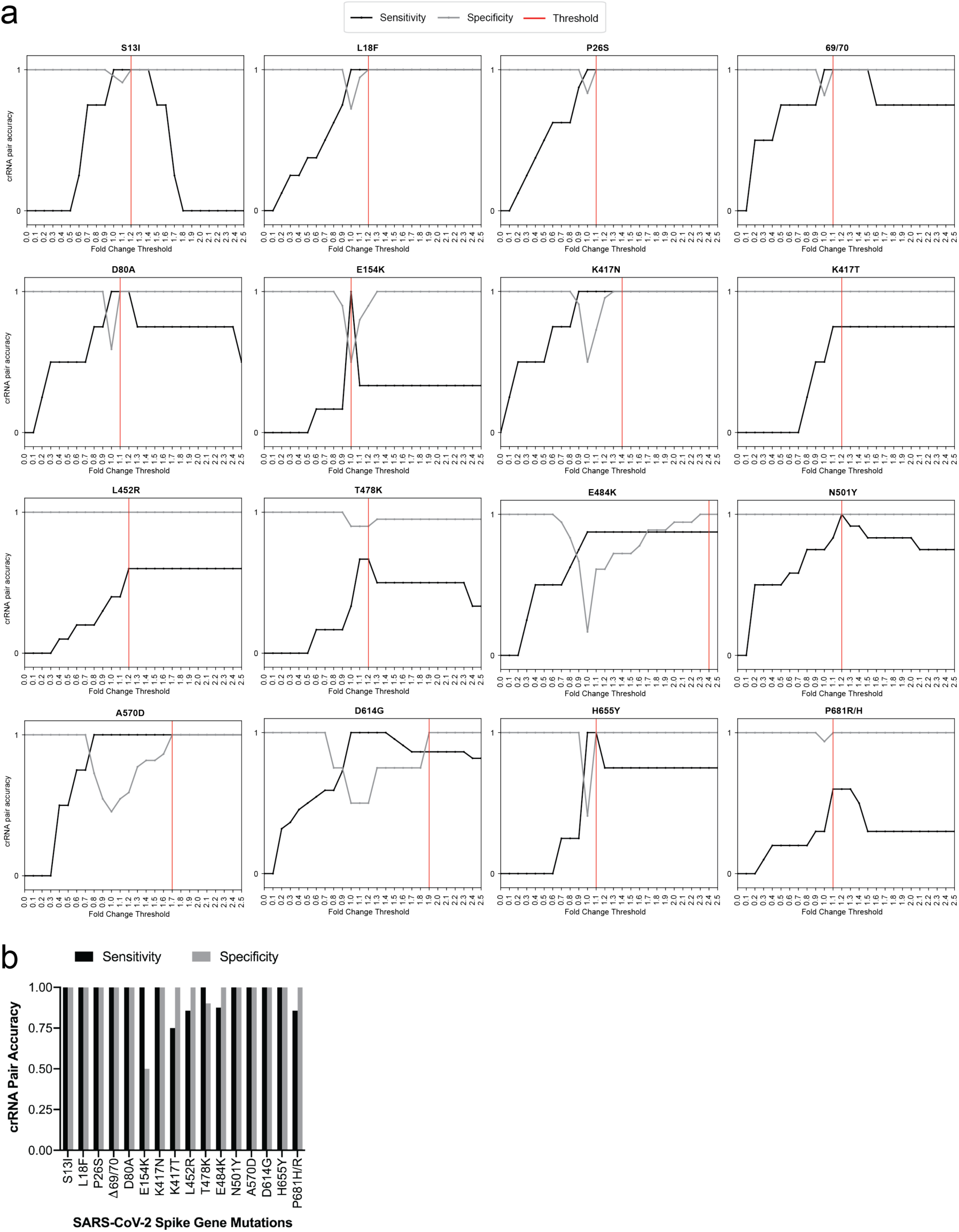
crRNA performance in Variant Identification Panel (VIP). **a,** Evaluation of sensitivity and specificity of crRNA pair detection to establish mutation calling threshold. All values based on viral seed stock testing. Black line: sensitivity/true positive rate; gray line: specificity/true negative rate; red line: mutation calling threshold. **b,** Summary of sensitivity and specificity results from **a**. Black: sensitivity; gray: specificity.

**Extended Data Figure 13.**
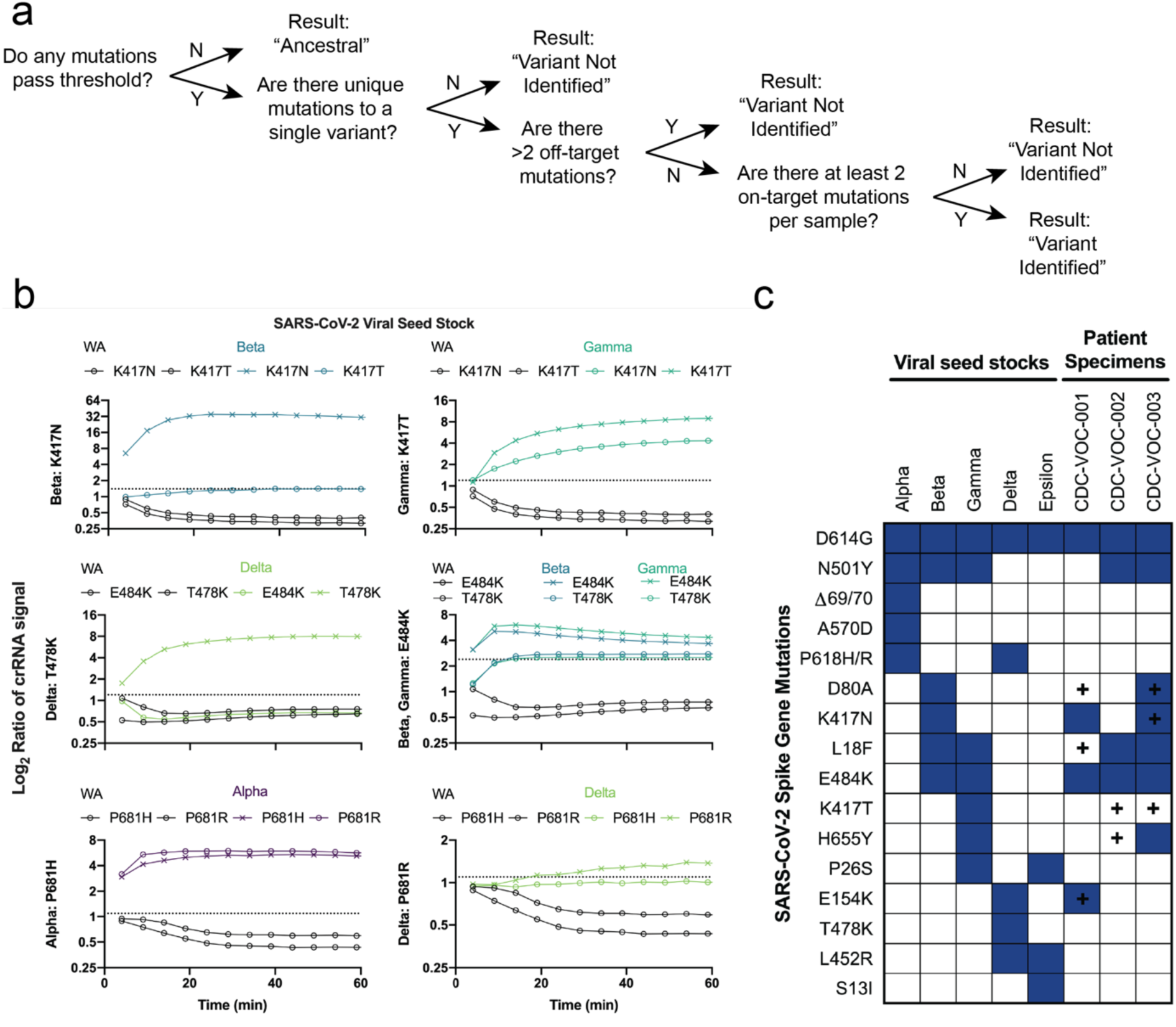
General variant identification analysis pipeline workflow and special cases. **a**, Variant identification analysis pipeline method for making variant calls based on fluorescence signals from VIP assay. **b**, Alpha, Beta, Gamma, and Delta viral seed stocks were evaluated using VIP. Kinetic curve data shown as the log_2_ of the maximum crRNA fluorescence ratio of mutation/ancestral at five min intervals for the first 60 min post-reaction initiation. Black: ancestral seed stock WA; purple: Alpha; blue: Beta; teal: Gamma; green: Delta. X lines represent the expected SNP based on NGS. O lines represent the unexpected SNP based on NGS. Dashed line represents threshold for variant calling based on Extended Data Fig. 12. **c**, Left, expected signal for the mutations that make up 5 SARS-CoV-2 variant lineages based on results from viral seed stock testing. Right, mutation calls from the variant identification analysis pipeline for three SARS-CoV-2 positive patient specimens where the analysis pipeline returns either a not identified result or an incorrectly identified variant result. Plus sign (+) represents signal that is either unexpected or expected for the given variant lineage based on NGS results.

**Extended Data Figure 14.**
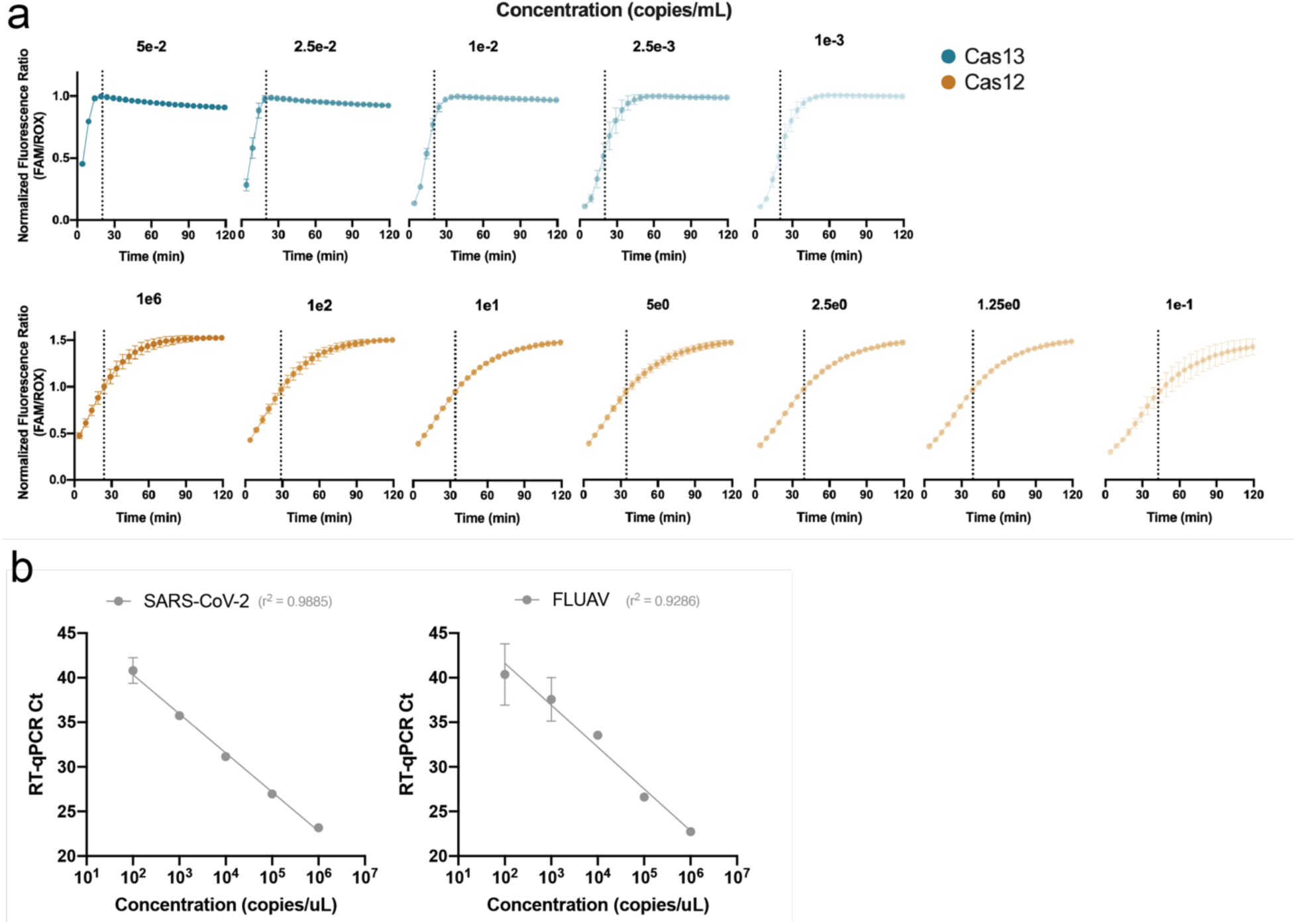
Sample quantification with combined Cas12 and Cas13 detection on mCARMEN RVP. **a,** Kinetic curves of SARS-CoV-2 detection by both Cas13 (blue) and Cas12 (orange) for 5 or 7 concentrations, respectively, up to 120 min. Dashed lines represent the time at which signal reaches 50% (IC_50_ infection point) for each concentration of target. **b**, Standard curves for SARS-CoV-2 and FLUAV based on RT-qPCR Ct values. Data related to Fig. 5c, now showing a standard 10-fold dilution series.

**Supplementary Table 1.** Cost calculation breakdown concerning mCARMEN.

| Category | Cost (USD) | Notes |
| --- | --- | --- |
| Fixed cost per chip | 568.7 | Includes reagents for Fluidigm and chip itself |
| Marginal cost per sample | 9.27 | Includes extraction, PCR reagents, and detection sample reagents |
| Marginal cost per virus tested | 1.1 | Includes CRISPR-Cas13a protein and assay reagents |

**Supplementary Table 2. Oligonucleotides related to this work.**

**Supplementary Table 3. Patient specimens related to this work.**

**Supplementary Table 4.** Preliminary limit of detection (LOD) results using synthetic targets from 1-10,000,000 copies/mL. A range finding study for LOD for each target was conducted using synthetic materials spiked into pooled negative patient specimens stored in Universal Transport Medium (UTM). The range finding study was conducted with a dilution series ranging from 1 - 10,000,000 copies/mL. The estimated LOD range was determined for each viral target between the lowest dilution detected and the highest undetected concentration for the confirmatory experiments.

| Target | SARS-CoV-2 |  |  |  |  |  |  |  |
| --- | --- | --- | --- | --- | --- | --- | --- | --- |
| copies/mL | 10,000,000 | 1,000,000 | 100,000 | 10,000 | 1,000 | 100 | 10 | 1 |
| Positive/total | 5/5 | 5/5 | 5/5 | 5/5 | 3/5 | 0/5 | 0/5 | 0/5 |

| Target | HCoV-HKU1 |  |  |  |  |  |  |  |
| --- | --- | --- | --- | --- | --- | --- | --- | --- |
| copies/mL | 10,000,000 | 1,000,000 | 100,000 | 10,000 | 1,000 | 100 | 10 | 1 |
| Positive/total | 5/5 | 5/5 | 5/5 | 4/5 | 2/5 | 0/5 | 0/5 | 0/5 |

| Target | HCoV-NL63 |  |  |  |  |  |  |  |
| --- | --- | --- | --- | --- | --- | --- | --- | --- |
| copies/mL | 10,000,000 | 1,000,000 | 100,000 | 10,000 | 1,000 | 100 | 10 | 1 |
| Positive/total | 5/5 | 5/5 | 5/5 | 5/5 | 5/5 | 3/5 | 0/5 | 0/5 |

| Target | HCoV-OC43 |  |  |  |  |  |  |  |
| --- | --- | --- | --- | --- | --- | --- | --- | --- |
| copies/mL | 10,000,000 | 1,000,000 | 100,000 | 10,000 | 1,000 | 100 | 10 | 1 |
| Positive/total | 5/5 | 5/5 | 5/5 | 4/5 | 2/5 | 2/5 | 0/5 | 0/5 |

| Target | FLUAV |  |  |  |  |  |  |  |
| --- | --- | --- | --- | --- | --- | --- | --- | --- |
| copies/mL | 10,000,000 | 1,000,000 | 100,000 | 10,000 | 1,000 | 100 | 10 | 1 |
| Positive/total | 5/5 | 5/5 | 5/5 | 3/5 | 2/5 | 0/5 | 0/5 | 0/5 |

| Target | FLUBV |  |  |  |  |  |  |  |
| --- | --- | --- | --- | --- | --- | --- | --- | --- |
| copies/mL | 10,000,000 | 1,000,000 | 100,000 | 10,000 | 1,000 | 100 | 10 | 1 |
| Positive/total | 5/5 | 5/5 | 5/5 | 5/5 | 5/5 | 1/5 | 0/5 | 0/5 |

| Target | HPIV-3 |  |  |  |  |  |  |  |
| --- | --- | --- | --- | --- | --- | --- | --- | --- |
| copies/mL | 10,000,000 | 1,000,000 | 100,000 | 10,000 | 1,000 | 100 | 10 | 1 |
| Positive/total | 5/5 | 5/5 | 5/5 | 5/5 | 5/5 | 1/5 | 0/5 | 0/5 |

| Target | HRSV |  |  |  |  |  |  |  |
| --- | --- | --- | --- | --- | --- | --- | --- | --- |
| copies/mL | 10,000,000 | 1,000,000 | 100,000 | 10,000 | 1,000 | 100 | 10 | 1 |
| Positive/total | 5/5 | 5/5 | 5/5 | 5/5 | 3/5 | 0/5 | 1/5 | 0/5 |

| Target | HMPV |  |  |  |  |  |  |  |
| --- | --- | --- | --- | --- | --- | --- | --- | --- |
| copies/mL | 10,000,000 | 1,000,000 | 100,000 | 10,000 | 1,000 | 100 | 10 | 1 |
| Positive/total | 5/5 | 5/5 | 5/5 | 4/5 | 1/5 | 0/5 | 0/5 | 0/5 |

**Supplementary Table 5.**
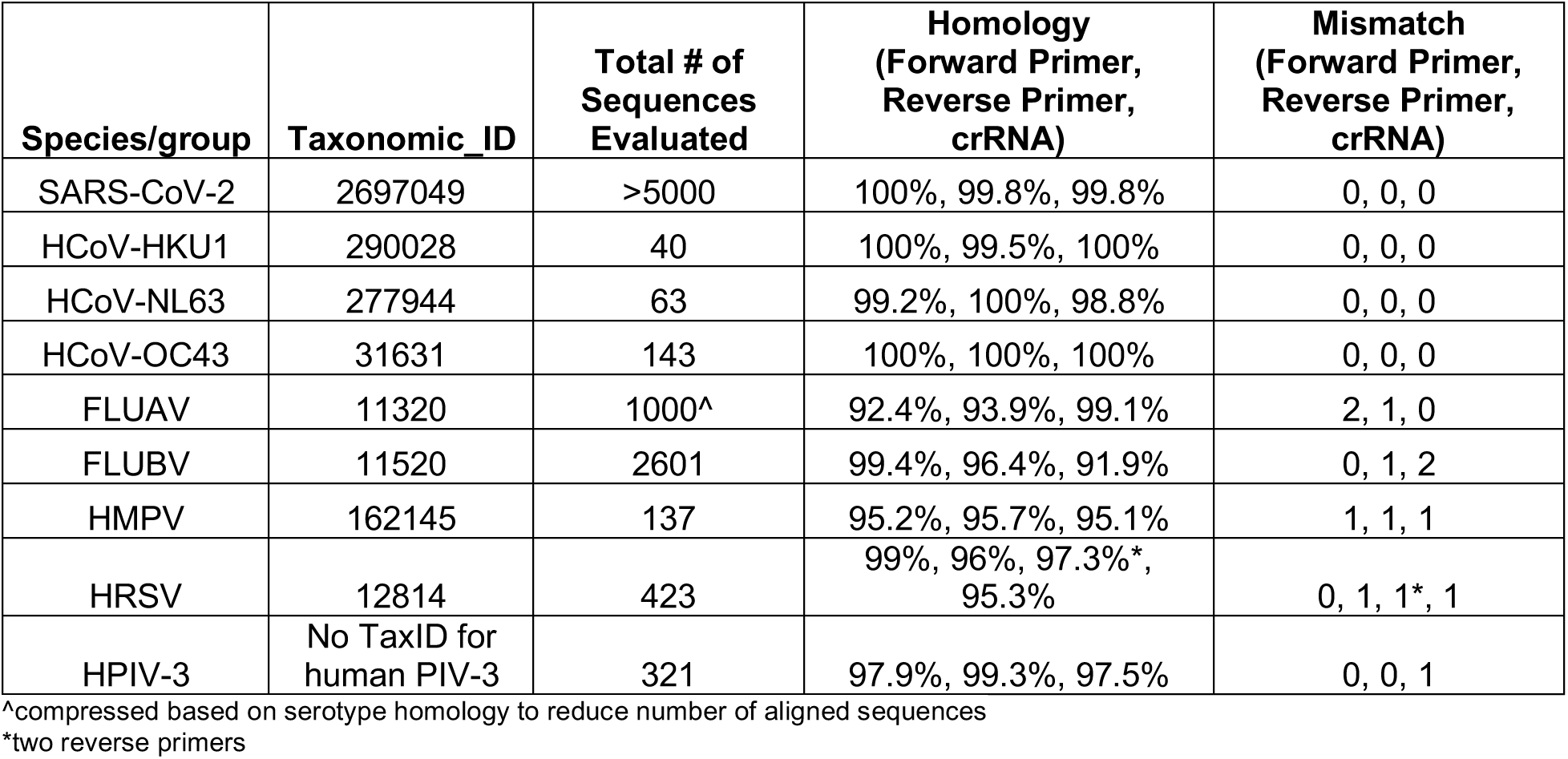
*In silico* inclusivity evaluation of CARMEN-RVP. Inclusivity was evaluated by performing an *in silico* analysis using all publicly available sequences of all targets on the panel. Inclusivity was tested by performing an *in silico* analysis using all publicly available sequences of all targets on the panel. Complete genomes for all viruses were downloaded from NCBI on April 2^nd^ 2021 and aligned using MAFFT, a multiple sequence alignment program that stands for “Multiple Alignment using Fast Fourier Transform.” MAFFT utilizes several different alignment algorithms. For viral species with less than 1000 sequences, the FFT-NS-ix1000 algorithm was used to create the MAFFT alignment. For viral species with >1000 sequences, the FFT-NS-1 algorithm was used to create the MAFFT alignment. The primer and crRNA sequences were then mapped to the aligned viral sequences using a consensus alignment to determine the percent identity (homology) and the number of mismatches. The average homology and mismatches were taken across the total number of sequences evaluated. Please note that mismatches below for crRNA sequences do not take wobble base pairing (G-U pairing) into account. Additionally the SARS-CoV-2 crRNA and primer sequences were compared by NCBI BLAST+ against the nr/nt databases (updated 03/31/2021, N=68965867 sequences analyzed) and the Betacoronavirus database (updated 04/01/2021, N=140760). The search parameters were adjusted to blastn-short for short input sequences. The match and mismatch scores are 1 and -3, respectively. The penalty to create and extend a gap in an alignment is 5 and 2, respectively. Blast results confirmed only perfect matches to SARS-CoV-2.

**Supplementary Table 6.** *In silico* Cross-reactivity (analytical specificity) evaluation of CARMEN-RVP. Complete genomes for all viruses were downloaded from NCBI on April 2nd 2021 and aligned using MAFFT. For viral species with less than 1000 sequences, FFT-NS-ix1000 was used. For viral species with >1000 sequences, FFT-NS-1 was used for the MAFFT alignment. The primer and crRNA sequences were then mapped to the aligned viral sequences using a consensus alignment to determine percent identity (homology). The average homology was taken across the panel sequences and the total number of sequences evaluated. Bolded text represents on-target primers/crRNA to the intended viral sequences. Not all sequence combinations were evaluated since whole genome homology between many viruses is significantly less than 80%. None of the primer or crRNA sequences has >80% homology to other, unintended viral or bacterial sequences, making the panel highly specific to the particular viruses of interest. More specifically, no *in silico* cross-reactivity >80% homology between any primers and crRNA sequences on the CARMEN-RVP assay is observed for the following common respiratory flora and other viral pathogens: SARS-CoV-1, HCoV-MERS, Adenovirus, Enterovirus, Rhinovirus, *Chlamydia pneumoniae, Haemophilus influenzae, Legionella pneumophila, Mycobacterium tuberculosis, Streptococcus pneumoniae, Streptococcus pyogenes, Bordetella pertussis, Mycoplasma pneumoniae, Pneumocystis jirovecii, Candida albicans, Pseudomonas aeruginosa, Staphylococcus epidermis, Streptococcus salivarius*.

|  | Panel Sequences (Forward primer, reverse primer, crRNA)<br>Percent homology |  |  |  |  |  |  |  |  |
| --- | --- | --- | --- | --- | --- | --- | --- | --- | --- |
| Homology Sequences | SARS-CoV-2 | HCoV-HKU1 | HCoV-NL63 | HCoV-OC43 | FLUAV | FLUBV | HMPV | HRSV | HPIV-3 |
| SARS-CoV-2 | **100%** | 63.30% | 60.80% | 61.80% | 60% | 64.40% | 66.70% | 54.60% | 66.90% |
| HCoV-229E | 65.40% | 62.60% | 72.10% | 68.50% |  |  |  |  |  |
| HCoV-HKU1 | 65.00% | **99.80%** | 63.50% | 74.50% |  |  |  |  |  |
| HCoV-NL63 | 60.40% | 62.90% | **99.30%** | 68.90% |  |  |  |  |  |
| HCoV-OC43 | 56% | 72.90% | 66.50% | **100%** |  |  |  |  |  |
| HCoV-MERS | 58.90% | 59.80% | 63.20% | 63.10% |  |  |  |  |  |
| SARS-CoV-1 | 62.90% | 61.00% | 53.90% | 67.50% |  |  |  |  |  |
| FLUAV | 60.40% |  |  |  | **95.80%** | 70.80% |  |  |  |
| FLUBV | 67.50% |  |  |  | 77% | **95.90%** |  |  |  |
| FLUCV | 57.50% |  |  |  | 69.50% | 63.90% |  |  |  |
| HMPV | 48.80% |  |  |  |  |  | **95.30%** | 71.90% | 63% |
| HRSV | 67.50% |  |  |  |  |  | 59.10% | **96.90%** | 56% |
| HPIV-3 | 68.30% |  |  |  |  |  | 67.10% | 61.50% | **98.20%** |
| HPIV-1 | 64.70% |  |  |  |  |  | 63.20% | 59.20% | 68.90% |
| HPIV-2 | 63.40% |  |  |  |  |  | 64.50% | 64.70% | 65% |
| HPIV-4 | 63.90% |  |  |  |  |  | 55.80% | 49% | 60% |
| HRV | 43.70% |  |  |  |  |  | 46.30% | 47.60% | 43.80% |
| Ad71 | 22.90% |  |  |  |  |  |  |  |  |
| EV-D68 | 60.90% |  |  |  |  |  |  |  |  |

**Supplementary Table 7.** Viral seed stock and genomic RNA information related to this work.

| Virus Name | Cat# | Description | Vendor | NGS Amino Acid Substitutions |
| --- | --- | --- | --- | --- |
| SARS-CoV-2 | VR-1986HK | Viral seed stock | ATCC | N/A |
| HPIV-3 | VR-93 | Viral seed stock | ATCC | N/A |
| OC43 | VR-1558 | Viral seed stock | BEI Resources | N/A |
| HRSV | VR-1540 | Viral seed stock | ATCC | N/A |
| FLUA(H3N2) | NR-43756 | Genomic RNA | BEI Resources | N/A |
| FLUBV | VR-1804 | Genomic RNA | BEI Resources | N/A |
| HMPV | NR-49122 | Genomic RNA | ATCC | N/A |
| SARS-CoV-2 Alpha | N/A | Viral seed stock | CDC | H69-, V70-, Y144-, N501Y, A570D, D614G, P681H, T716I, A771X, S982A, D1118H |
| SARS-CoV-2 Beta | N/A | Viral seed stock | CDC | L18F, D80A, D215G, T240~, L241-, L242-, A243~, K417N, E484K, N501Y, D614G, A701V, A771X |
| SARS-CoV-2 Gamma | N/A | Viral seed stock | CDC | L18F, T20N, P26S, D138Y, R190S, K417T, E484K, N501Y, D614G, H655Y, A771X, T1027I, R1039X, V1176F |
| SARS-CoV-2 Delta | N/A | Viral seed stock | CDC | T19R, G142D, E156-, F157-, R158G, L452R, T478K, D614G, P681R, D950N |
| SARS-CoV-2 Epsilon | N/A | Viral seed stock | CDC | S13I, P26S, W152C, L452R, D614G, A771X, R1039X |

## Notes

### Author Declarations

Use of clinical excess of human specimens from patients with SARS-CoV-2 from the Broad Institute Genomics Platform CLIA Laboratory was approved by the MIT IRB Protocol #1612793224. Additional SARS-CoV-2 samples were collected from consented individuals under Harvard Longwood Campus IRB #20-1877 and covered by an exempt determination (EX-7295) at the Broad Institute. Other human-derived samples from patients with SARS-CoV-2 were collected by the CDC and determined to be non-human subjects research; the Broad Office of Research Subject Protections determined these samples to be exempt. Human specimens from patients with SARS-CoV-2, HCoV-HKU1, HCoV-NL63, FLUAV, FLUBV, HRSV, and HMPV were obtained under a waiver of consent from the Mass General Brigham IRB Protocol #2019P003305. Researchers at Princeton were determined to be conducting not-engaged human subjects research by the Princeton University IRB.

